# What Matters Most: A Multi-Stakeholder Study of Outcome Domains in Lower-Limb Prosthesis Use

**DOI:** 10.64898/2026.08.31.26361544

**Authors:** Mustafa Ahmed, Steven Karlsson-Brown, Pelagia Koufaki, Moghadaseh Ahmadi, Encarna M. Micó-Amigo

**Author notes:** **Corresponding author:** Encarna M. Micó-Amigo.

## Abstract

**Purpose:** Lower-limb prosthesis use involves interacting physical, psychosocial, and device-related outcomes that may not be fully represented by conventional clinical assessment. This study aimed to develop and evaluate a stakeholder-informed framework of outcome domains relevant to meaningful everyday prosthesis use.

**Materials and Methods:** A mixed-methods participatory design comprised synthesis of selected clinically relevant content from five established patient-reported outcome measures; semi-structured interviews and importance and actionability ratings with 18 contributors (12 prosthesis users, four clinicians, and two industrial partners); and integration of synthesis, qualitative, and rating findings. Interview records were analysed using reflexive thematic analysis and ratings descriptively.

**Results:** The resulting framework comprised four interrelated domains: Mobility, Physical Function, Psychosocial Wellbeing, and Prosthesis Experience. Mobility showed the clearest convergence across stakeholder perspectives. Prosthesis users showed the largest importance–actionability gap for Prosthesis Experience (4.5 vs 3.0), whereas clinicians showed the largest gap for Psychosocial Wellbeing (5.0 vs 3.0). Interviews highlighted day-to-day variability and the influence of confidence, fatigue, comfort, environment, social context, and device usability.

**Conclusions:** Meaningful outcome assessment in prosthetic rehabilitation should extend beyond mobility alone to consider physical function, psychosocial wellbeing, and prosthesis experience within everyday contexts. The framework provides a stakeholder-informed foundation for multidimensional outcome assessment.

## 1 Introduction

Adults with lower-limb amputation often require long-term, adaptive prosthetic care, as their functional abilities, personal goals, and environmental demands evolve over time (Dupuis et al., 2025). Prosthesis users value mobility, participation, self-efficacy, acceptance, socket comfort, and perceived safety (McDonald et al., 2019; Ostler et al., 2024b; Schaffalitzky et al., 2011). Satisfaction is multidimensional and biopsychosocial, shaped by appearance, weight, noise, socket fit and comfort, residual-limb health, and context of use (Luza et al., 2020; Baars et al., 2018). Users prioritise a range of outcomes, including participation, independence, confidence in public, perceived safety, socket comfort, and acceptance (Alluhydan et al., 2023; Diment et al., 2022), though the relative importance of these priorities varies across individuals and evolves across the recovery trajectory (Sanders et al., 2020). To ensure appropriate care provision, clinical assessment must therefore extend beyond physical impairment and device functionality, and consider the wider personal and environmental context, including the development of the prosthesis– user relationship, the user’s capacity, performance, functioning, physical activity, social participation, mental health, and overall quality of life (Dupuis et al., 2025; Schaffalitzky et al., 2012).

Outcome measurement may capture a range of domains relevant to prosthesis use, including mobility, physical function, pain, participation, emotional well-being, satisfaction, and quality of life (Heinemann et al., 2014; Ostler et al., 2023). By quantifying functional status and identifying minimal clinically important changes that exceed measurement error, outcome measurement can guide rehabilitation services such as socket or component adjustments and rehabilitation planning (Heinemann et al., 2014; Walton et al., 2015). In prosthetic practice, two broad approaches are used: patient-reported outcome measures (PROMs) and performance-based or observed measures (Ostler et al., 2022; Heinemann et al., 2014). PROMs capture the lived experience of prosthesis users and include instruments such as the Prosthesis Evaluation Questionnaire, the Trinity Amputation and Prosthesis Experience Scales, and the Prosthetic Limb Users Survey of Mobility (Legro et al., 1998; Gallagher and MacLachlan, 2000; Hafner et al., 2017). Observed performance measures assess functional ability under standardised conditions, including the Timed Up and Go, the Six-Minute Walk Test, and the Amputee Mobility Predictor (Condie et al., 2006; Hawkins and Riddick, 2018).

Standardised PROMs and performance-based measures are not consistently integrated into routine prosthetic practice, with time, space, and administration requirements presenting barriers to their use (Morgan et al., 2022). Moreover, clinic-based performance measures primarily assess what an individual can do under standardised conditions (capacity), which may not fully reflect what they actually do in everyday life (performance) (Parker et al., 2010). In particular, limb-loss-specific instruments may not fully capture external factors such as uneven terrain, obstacles, external loads, lighting, crowds, and weather conditions, or internal factors such as fatigue, confidence, and pain, all of which can influence prosthesis use in daily life (Chadwell et al., 2020; Day et al., 2019; Wurdeman et al., 2018). Patients have also reported that outcome measures used in routine prosthetic care do not always capture an accurate or meaningful picture of recovery, particularly when standardised metrics do not align with priorities in day-to-day life (Ostler et al., 2024b). Their routine implementation is further constrained by time pressure, workflow and space limitations, training gaps, and uncertainty regarding the selection of appropriate tools and outcomes (Gaunaurd et al., 2015; Morgan et al., 2022; Young et al., 2018). These limitations may contribute to misalignment between what is measured and what matters to prosthesis users, while also limiting consistency and comparability across prosthetic services (Condie et al., 2006).

Patient and Public Involvement (PPI) refers to research carried out with or by members of the public rather than to, about, or for them (National Institute for Health and Care Research, 2021). PPI can improve the relevance and impact of health research by incorporating lived experience and stakeholder priorities (Brett et al., 2014). Consensus initiatives, including the Core Outcome Measures in Effectiveness Trials (COMET) Initiative and the International Consortium for Health Outcomes Measurement (ICHOM), emphasise stakeholder involvement in identifying and prioritising outcomes before selecting appropriate measurement instruments (Williamson et al., 2017; International Consortium for Health Outcomes Measurement, nd). The International Classification of Functioning (ICF) offers a useful scaffold across body functions, activities, participation, and context, but deductive mappings can miss prosthesis-specific concepts such as appearance, confidence, and socket comfort, which justifies inductive and PPI-led approaches (McDonald et al., 2019; Schaffalitzky et al., 2011).

Prior work has advanced understanding of meaningful outcomes following lower-limb amputation, but important gaps remain. Some studies have focused on outcomes associated with prosthetic prescription or specific prosthetic components (Schaffalitzky et al., 2011; McDonald et al., 2019), while others have identified patient-valued outcome domains and examined patients’ experiences of outcome measurement (Ostler et al., 2023, 2024b). Contextual modifiers of everyday performance, including terrain, obstacles, environmental conditions, fatigue, and pain, are also recognised as important influences on prosthesis use (Chadwell et al., 2020; Day et al., 2019). An important remaining question is how concepts represented in established outcome measures align with priorities identified by prosthesis users, clinicians, and industry within a common framework. Although patient-valued domains and a range of patient-reported and performance-based measures have been described, the relationship between concepts represented in these measures and stakeholder-valued outcomes remains insufficiently characterised (Condie et al., 2006; Ostler et al., 2024a; Hawkins and Riddick, 2018).

The study addressed these gaps through a preparatory co-design phase followed by three sequential, linked steps. The preparatory PPI and stakeholder co-design phase involved prosthesis users, clinicians, and an industrial representative in refining the study design, outcome-measure selection approach, data collection procedures, rating exercise, plainlanguage materials, and recruitment strategy before the main evaluation phase. First, a structured, concept-focused synthesis of selected clinically relevant content from established PROMs was undertaken to develop a preliminary domain–theme framework. Content selection was purposive and non-exhaustive, with the aim of identifying clinically meaningful concepts relevant to prosthesis use, everyday functioning, and rehabilitation rather than comprehensively analysing every item within each instrument. Second, the resulting framework was evaluated and refined through semi-structured interviews with prosthesis users, clinicians, and industrial partners, who explored the relevance and meaning of the proposed domains in everyday prosthesis use and rated each domain for importance and actionability. Third, the selected PROM-content synthesis, qualitative interview findings, and quantitative ratings were integrated across stakeholder groups to identify areas of convergence and divergence and to compare stakeholder priorities with patterns represented within the selected PROM content. These strands were subsequently synthesised into a stakeholder-informed, context-sensitive four-domain framework, recognising that everyday prosthesis use may be influenced by internal factors such as fatigue, pain, mood, and task demands, as well as external factors such as terrain, weather, obstacles, lighting, and social context.

## 2 Methods

### 2.1 Study design and reporting framework

This study employed a mixed-methods, participatory design comprising a preparatory PPI and stakeholder co-design phase followed by three sequential, linked study steps. The preparatory phase was undertaken to refine the study design, outcome-measure selection approach, data collection procedures, rating exercise, plain-language materials, and recruitment strategy before the main evaluation phase. The three subsequent steps comprised: (1) development of a structured framework of domains through literature synthesis and mapping of selected clinically and conceptually relevant content from established patient-reported outcome measures (PROMs); (2) evaluation and refinement of the resulting framework through semi-structured interviews with prosthesis users, clinicians, and industrial partners; and (3) integrative analysis combining qualitative insights with structured domain ratings across stakeholder groups.

Reporting of PPI followed the GRIPP2 guidance (Staniszewska et al., 2017), and the qualitative components were described in accordance with the Standards for Reporting Qualitative Research (SRQR) (O’Brien et al., 2014). The qualitative strand was informed by an interpretivist stance, recognising that meanings attached to recovery and prosthesis use are socially situated and context dependent (Lincoln and Guba, 1985). Throughout the study, individuals providing lived, clinical, or industrial perspectives are referred to as contributors, reflecting the participatory orientation of the work. In the main framework-evaluation phase, contributors’ interview accounts and structured ratings additionally constituted qualitative and quantitative study data.

### 2.2 Preparatory PPI and stakeholder co-design phase

#### 2.2.1 Co-design contributors and procedure

Before finalisation of the main study procedures and materials, a preparatory co-design phase was conducted with seven contributors representing lived, clinical, and industrial perspectives. The group comprised four prosthesis users, two clinicians, and one industrial representative. The co-design phase was distinct from the subsequent framework-evaluation phase. Six of the seven co-design contributors did not participate in the main evaluation phase; one clinician contributed to both phases. The co-design activities focused on study procedures, materials, outcome-measure selection, recruitment, and the design of the rating exercise rather than on rating or prioritising the final outcome domains.

Co-design contributors participated in one-to-one, semi-structured online sessions lasting approximately 45–60 minutes. Sessions were conducted using Microsoft Teams or Google Meet according to contributor preference. Discussion focused on four areas: (1) questionnaire selection and content; (2) proposed data collection procedures; (3) accessibility and clarity of the plain-language study materials; and (4) recruitment and engagement strategies intended to support inclusive participation. Contemporaneous researcher notes were taken during and immediately after the co-design sessions and were used to populate the study change log.

Contributors reviewed the preliminary set of outcome measures identified through the initial literature work, including the Prosthesis Evaluation Questionnaire (PEQ), Trinity Amputation and Prosthesis Experience Scales (TAPES), Prosthetic Limb Users Survey of Mobility (PLUS-M), Orthotics and Prosthetics User Survey (OPUS), and Locomotor Capabilities Index-5 (LCI-5). They were asked to consider the relevance of the constructs represented, areas that appeared insufficiently captured, and practical considerations such as questionnaire length, administration burden, and scoring complexity.

Contributors also reviewed the proposed semi-structured interview format, the five-point domain-rating exercise, and the planned approach to documenting interviews. Draft plain-language descriptions of the study, proposed domains, and rating exercise were reviewed for clarity and accessibility. Contributors were additionally invited to comment on the proposed recruitment approach and recommend strategies for improving reach and inclusivity. Participation was supported through accessible materials, flexible scheduling, and inclusive communication. PPI contributors were remunerated in accordance with National Institute for Health and Care Research (NIHR) guidance.

#### 2.2.2 Integration of co-design input

Co-design input was reviewed using a pragmatic, theme-based approach focused on identifying actionable recommendations for refining the subsequent study procedures and materials. A change log was maintained to document each recommendation, the decision made in response, and the component of the study affected.

The co-design phase resulted in several substantive refinements. All five identified outcome measures were retained as source instruments for the structured synthesis, while selected content considered most relevant to prosthesis use, everyday functioning, and rehabilitation was taken forward for mapping rather than administering the complete instruments to contributors. Feedback also informed the inclusion and refinement of concepts such as prosthesis usability, including donning and doffing, and environmental influences on mobility, including terrain, weather, and social context.

Plain-language terminology was revised in response to contributor feedback. This included adoption of the term “prosthesis experience” and clearer explanations of mobility and psychosocial wellbeing. Recruitment procedures were broadened to include communitybased and user-led routes, while provision for language support was incorporated to facilitate participation where required.

The distinction between domain importance and actionability was also clarified during the co-design process. Actionability was retained as a study construct and operationalised in the main evaluation phase as the perceived practicality of assessing or monitoring a domain within regular care. In this study, actionability therefore referred specifically to practical assessability within routine care rather than to the broader concept of whether an outcome could directly trigger a clinical action. Plain-language explanations and examples were incorporated into the rating materials to support consistent interpretation.

### 2.3 Ethics

The study received approval from the School of Engineering and Physical Sciences, Heriot-Watt University Research Ethics Committee (Ref: 2025-11249-15247) and was conducted in accordance with the principles of the Declaration of Helsinki. Recruitment for the main framework-evaluation phase was conducted between June and September 2025. All participants in the main framework-evaluation phase received appropriate study information and provided written informed consent, with the right to withdraw at any point.

Framework-evaluation interviews were audio-recorded in accordance with the approved ethical protocol to support accurate documentation of contributor accounts, qualitative analysis, and verification of direct quotations. Recordings were retained throughout data preparation and analysis and were deleted only after the qualitative analysis and quotation verification had been completed, in accordance with the approved data-management procedure. Study records were pseudonymised for analysis and reporting and stored securely on password-protected, university-managed systems in accordance with institutional policies and the UK General Data Protection Regulation (UK GDPR).

### 2.4 Step 1: Literature synthesis and framework development

The first study step focused on a structured, concept-focused synthesis of outcome measures used in lower-limb prosthetic rehabilitation. Instruments were considered if they were developed or validated for prosthesis users, were widely used or recognised in prosthetic rehabilitation research, assessed constructs relevant to prosthesis use and rehabilitation, were available in English, and reported psychometric properties such as reliability, validity, or responsiveness (Mokkink et al., 2010). PROMs were the primary focus of the mapping. The synthesis was intended to support development of a clinically relevant domain framework rather than to provide an exhaustive item-by-item content analysis of each instrument.

Relevant measures were identified in January 2025 through targeted searches of PubMed, Scopus, and Web of Science, supplemented by existing reviews and citation tracking. The purpose of this search was to identify established PROMs relevant to lower-limb prosthetic rehabilitation rather than to undertake a systematic review of all available outcome measures. The preliminary set of measures was identified by a co-author and subsequently reviewed during the preparatory co-design phase, where contributors considered the relevance of the constructs represented, areas that appeared insufficiently captured, and practical characteristics such as questionnaire length, administration burden, and scoring complexity. The final set comprised the Prosthesis Evaluation Questionnaire (PEQ) (Legro et al., 1998), the Trinity Amputation and Prosthesis Experience Scales (TAPES) (Gallagher and MacLachlan, 2000), the Prosthetic Limb Users Survey of Mobility (PLUS-M) (Hafner et al., 2017), the Orthotics and Prosthetics User Survey (OPUS) (Heinemann et al., 2003), and the Locomotor Capabilities Index-5 (LCI-5) (Franchignoni et al., 2004). The main characteristics and focus areas of these instruments are summarised in Table 1. Following co-design feedback regarding the burden and practical limitations of administering multiple complete instruments, the measures were used as source instruments for a selected-content, concept-focused synthesis rather than administered as questionnaires to contributors.

**Table 1:** PROMs included in the synthesis and their main characteristics.

| Instrument | Core constructs/domains targeted (as defined by the PROM) | Response Format |
| --- | --- | --- |
| **PEQ** (Prosthesis Evaluation Questionnaire) | Satisfaction, mobility, appearance, residual-limb health, social burden | Visual analogue scale (VAS): continuous, 0 (most negative) to 100 (most positive) |
| **TAPES** (Trinity Amputation and Prosthesis Experience Scales) | Adjustment, satisfaction, activity restriction, psychosocial impact | Limitation scale: 3-point (1 = not at all limited; 3 = limited a lot). Psychosocial scales: 5-point Likert (1 = strongly disagree; 5 = strongly agree) |
| **PLUS-M** (Prosthetic Limb Users Survey of Mobility) | Perceived mobility across varied environments | Difficulty scale: 5-point (1 = unable to do; 5 = without any difficulty) |
| **OPUS** (Orthotics and Prosthetics User Survey) | Function, quality of life, satisfaction with device and clinical services | Difficulty scale: 5-point (1 = unable to do; 5 = without any difficulty) |
| **LCI-5** (Locomotor Capabilities Index) | Mobility tasks from basic to advanced | Ordinal ability scale: 5-point (0 = unable; 4 = able without any aid); max score = 56 |

For each instrument, the lead researcher purposively selected items and content elements considered most relevant to prosthesis use, everyday functioning, and rehabilitation. Selected content was entered into a structured database capturing: (1) the source PROM; (2) the item or content represented; (3) the original subscale or conceptual area, where applicable; (4) the assigned outcome domain; and (5) the assigned theme or subtheme. Selection was concept-focused and deliberately non-exhaustive, with the aim of capturing clinically meaningful content capable of informing development of the preliminary framework rather than reproducing or analysing every item from each instrument. The resulting mapping therefore represented an analytic sample of relevant PROM content rather than a comprehensive content analysis of the complete instruments.

The selected content was coded and mapped collaboratively by the lead researcher and a second researcher. Coding was informed by the thematic analysis approach described by Braun and Clarke (2006, 2022). The researchers familiarised themselves with the selected content, assigned initial conceptual codes, compared areas of similarity and overlap, and iteratively grouped related concepts into higher-order themes and domains. Differences in interpretation were discussed during the mapping process and resolved through collaborative review and refinement of the developing framework. This process was intended to support conceptual organisation rather than to assess inter-rater reliability. The International Classification of Functioning, Disability and Health (ICF) (World Health Organization, 2001) served as a deductive scaffold, with identified concepts mapped to relevant ICF areas, including body functions, activities, and participation, where appropriate. Methodological guidance from the COMET Initiative (Williamson et al., 2017) and the International Consortium for Health Outcomes Measurement (ICHOM) (International Consortium for Health Outcomes Measurement, nd) informed the structured definition and organisation of outcome domains, including the use of standardised terminology and consideration of stakeholder-relevant outcomes.

The grouped categories were consolidated into four overarching domains with associated subthemes and illustrative examples (Table 2). Mobility was defined specifically as locomotion-related activities, including walking and terrain negotiation, reflecting its central role in prosthetic rehabilitation and its frequent assessment in clinical mobility measures (Hafner et al., 2017; Franchignoni et al., 2004). Physical function captured broader daily activities beyond locomotion, including activities of daily living and participation in routine tasks, thereby representing functional independence in everyday contexts (Heinemann et al., 2003; Gallagher and MacLachlan, 2000). Psychosocial wellbeing encompassed emotional, psychological, and social dimensions influencing adaptation and participation, while prosthesis experience addressed aspects related to comfort, usability, and interaction with the device and associated services. This structure was adopted to reduce conceptual overlap while remaining conceptually aligned with the ICF framework. Terminology and plain-language descriptions of the domains were subsequently refined through the preparatory co-design process before presentation in the main evaluation phase.

**Table 2:**
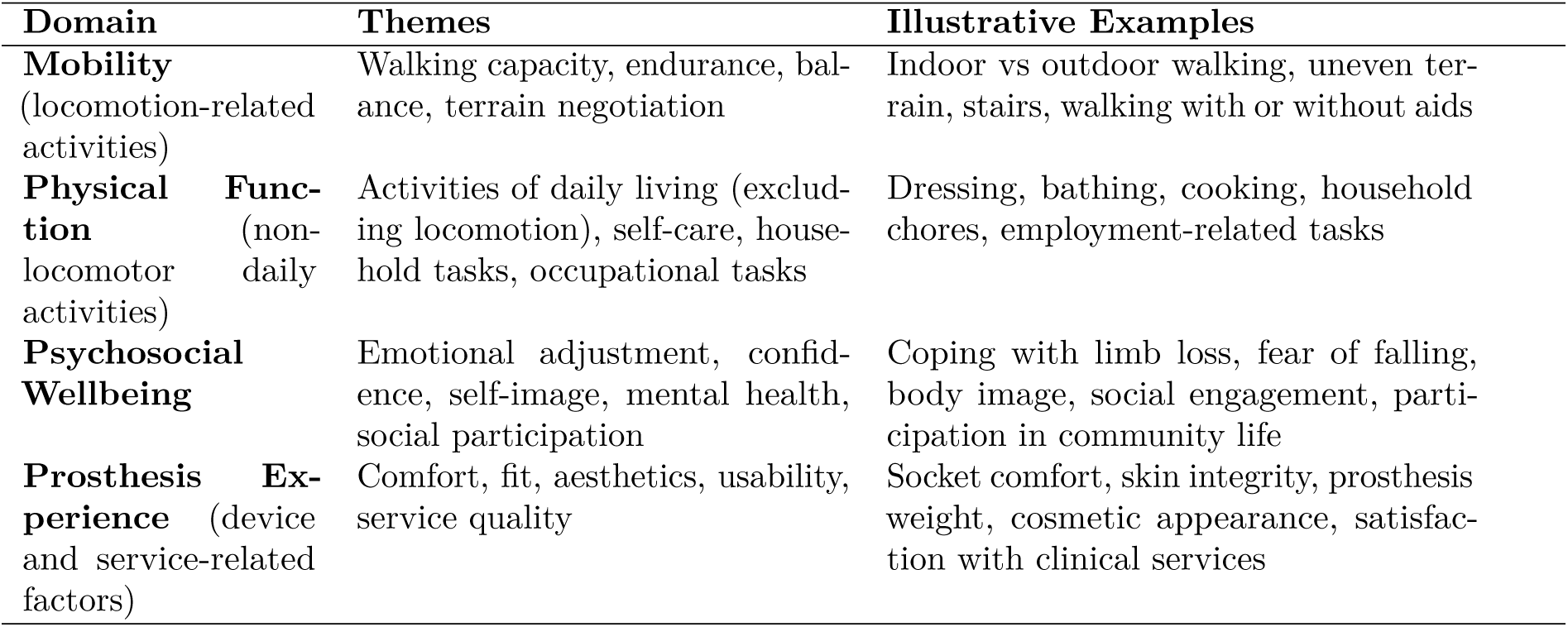
Overarching domains, subthemes, and illustrative examples.

### 2.5 Step 2: Contributor interviews and domain evaluation

#### 2.5.1 Contributors and recruitment

To evaluate and refine the domain framework, three stakeholder groups were engaged: prosthesis users, clinicians, and industrial representatives. In total, 18 contributors participated in the main evaluation phase, comprising twelve prosthesis users, four clinicians, and two industrial representatives.

Prosthesis users were eligible if they were aged 18 years or older and had at least two years of continuous prosthesis use. Participation was supported in English or, where required, through appropriate language support. Clinicians were eligible if they held clinical, rehabilitation, or clinically relevant research roles involving expertise in lower-limb prosthetic rehabilitation. Industrial representatives were eligible if they held a professional role in prosthetic device design, development, or sales, with a minimum of two years of industry experience.

#### 2.5.2 Equality, diversity and inclusion

Equality, diversity, and inclusion were formally embedded in the study design, reflecting concerns that rehabilitation research may underrepresent marginalised and minoritised communities and that their priorities may not be adequately represented in outcome development (Quigley et al., 2023; Kokorelias et al., 2025). Recruitment was conducted in partnership with the British Association of Prosthetists and Orthotists (BAPO) and community-based organisations to broaden participation beyond conventional clinical recruitment pathways, including engagement with organisations supporting refugees, asylum seekers, and individuals from minority ethnic communities.

The resulting sample included participants from a range of demographic and socioeconomic backgrounds. Six of the twelve prosthesis users identified as Black African or Arab, and three participants were from refugee or asylum-seeking backgrounds living in shared accommodation. Language accessibility was prioritised to reduce barriers to participation. One Arabic-speaking contributor and one Persian-speaking contributor required language support during the study. Arabic-language support was provided by the lead researcher, and Persian-language support was provided by a co-author. This support enabled contributors to express their experiences and respond to interview questions in their preferred language where required. Relevant interview content was subsequently represented in English for analysis and reporting. Accessible materials, flexible scheduling, and inclusive communication approaches were also employed throughout to support participation across differing levels of health literacy and digital access.

This approach was informed by concerns that rehabilitation research may overlook individuals at greater risk of exclusion and that outcome frameworks developed without diverse perspectives may inadequately represent the experiences and priorities of the populations they are intended to serve (Quigley et al., 2023; Manz et al., 2022).

#### 2.5.3 Interview procedure

Contributors took part in one-to-one, semi-structured interviews lasting approximately 45–60 minutes. Sessions were conducted online using Microsoft Teams, Google Meet, or Zoom according to contributor preference. A structured discussion guide ensured that all contributors reflected on the same four domains (Table 2), while follow-up prompts encouraged elaboration on daily variability, contextual influences such as terrain or fatigue, and the personal meaning of independence and activity.

Prosthesis users were encouraged to describe examples from everyday life and to reflect on the emotional, environmental, and social factors that shaped their prosthesis use. Clinicians and industrial partners were similarly encouraged to draw on their professional experience of prosthetic rehabilitation, device development, service provision, and interaction with prosthesis users. The semi-structured format allowed common areas to be explored across stakeholder groups while retaining sufficient flexibility to capture group-specific perspectives and experiences.

#### 2.5.4 Presentation of domains and rating exercise

Following the qualitative discussion, contributors completed structured ratings of the same four domains: Mobility, Physical Function, Psychosocial Wellbeing, and Prosthesis Experience. Plain-language and visually accessible materials refined during the preparatory co-design phase were used to support consistent understanding of the domains and rating exercise.

Each domain was rated according to two constructs: perceived importance and perceived actionability. Importance represented the extent to which the domain was considered relevant to everyday life and rehabilitation, while actionability was operationalised as the perceived practicality of assessing or monitoring the domain within regular care. Both constructs were rated using five-point Likert scales. Importance was rated from 1 (not at all important) to 5 (extremely important), and actionability from 1 (not at all practical to assess or monitor in regular care) to 5 (extremely practical).

Before completing the ratings, the interviewer explained each domain using straightforward examples, for instance describing mobility in terms of leaving the home independently and navigating everyday environments, or physical function in terms of activities such as cooking, cleaning, and managing household tasks. Clarification was provided where required to support consistent interpretation of the domains and rating constructs. Following the rating exercise, an open discussion explored the reasons underlying the scores, potential overlaps or omissions, and examples of how each domain was expressed in everyday life, clinical practice, or prosthetic design and service contexts.

#### 2.5.5 Data handling

Framework-evaluation interviews were audio-recorded and documented through detailed researcher-led notes and structured forms capturing both numerical ratings and explanatory comments. Audio recordings were used throughout data preparation and analysis to support accurate documentation and interpretation of contributor accounts and verification of direct quotations. Notes were elaborated and contextualised immediately after each interview while recollections remained fresh. Recordings were deleted only after the qualitative analysis and quotation verification had been completed, in accordance with the approved ethical and data-management procedures.

Quantitative responses were entered into a secure database for descriptive analysis. Narrative interview records were pseudonymised and prepared for thematic interpretation. Each main-study contributor was assigned a unique alphanumeric identifier corresponding to their stakeholder group (U for prosthesis user, C for clinician, and IP for industrial partner).

### 2.6 Step 3: Analysis

#### 2.6.1 Interview analysis

Interview records were analysed using reflexive thematic analysis informed by Braun and Clarke (2006, 2022). Initial coding was guided by the four predefined domains while remaining open to inductive insights, including concepts that extended beyond or challenged the preliminary framework. Codes and developing patterns were examined across contributors to identify shared experiences as well as differences between stakeholder groups.

A second analyst (co-author E.M., with extensive experience in qualitative research) reviewed a subset of pseudonymised interview records and the evolving thematic structure. This review was used as a process of reflexive dialogue and critical interpretation rather than as a test of inter-coder reliability. Alternative interpretations, deviant or negative cases, and the relationships between emerging themes were discussed to challenge assumptions and iteratively refine the thematic interpretation.

The analysis focused on lived and professional experiences, contextual influences, and perceived interconnections between physical, psychosocial, environmental, and prosthesis-related aspects of everyday life and rehabilitation.

#### 2.6.2 Quantitative analysis

Quantitative data from the domain-rating exercise were analysed descriptively for prosthesis users, clinicians, and industrial partners. For prosthesis users and clinicians, ratings of importance and actionability were summarised using medians and interquartile ranges (IQRs) as the primary descriptive statistics, reflecting the ordinal five-point response scale and the small stakeholder-group sample sizes. Means and standard deviations were additionally calculated to provide complementary descriptive information. Because only two industrial partners participated, their ratings were reported using medians and observed ranges rather than interpolated interquartile ranges.

Individual response distributions were examined to characterise the degree of agreement and variability within each stakeholder group. For each domain, the descriptive difference between median importance and median actionability ratings was also calculated as importance minus actionability to identify areas in which perceived relevance and perceived practicality of assessment diverged. No inferential statistical testing was undertaken because the quantitative component was exploratory and the stakeholder groups, particularly the clinician and industrial samples, were small. Quantitative findings were therefore interpreted descriptively as indicators of stakeholder priorities and the perceived practicality of assessing or monitoring each domain within regular care.

#### 2.6.3 Integration of findings

The selected PROM-content synthesis, qualitative interview findings, and structured domain ratings were integrated through a mixed-methods convergence approach. Patterns identified within the selected PROM content provided information about how clinically relevant concepts were represented across the source measures, quantitative summaries indicated the relative importance and perceived actionability of each domain, and qualitative accounts provided explanatory context regarding why particular outcomes were valued and how they were experienced in everyday life, clinical practice, and prosthetic design.

Findings were compared across prosthesis users, clinicians, and industrial partners to identify areas of convergence and divergence between stakeholder perspectives and across the three analytic strands. Integration focused on whether concepts represented within the selected PROM content were also emphasised by stakeholders, whether stakeholder accounts extended or contextualised those concepts, and where perceived importance differed from the perceived practicality of routine assessment or monitoring.

The integrated analysis was used to refine interpretation of the four-domain framework and to examine how Mobility, Physical Function, Psychosocial Wellbeing, and Prosthesis Experience interacted with internal and external contextual influences in everyday prosthesis use. This process informed the final context-aware framework. Because the PROM synthesis was purposive and non-exhaustive, comparisons with stakeholder findings were interpreted as patterns within the selected content rather than as evidence of complete coverage or omission within the source instruments.

## 3 Results

### 3.1 Preparatory PPI and Stakeholder Co-design Outcomes

Seven contributors participated in the preparatory co-design phase, comprising four prosthesis users, two clinicians, and one industrial representative. Six of these contributors did not participate in the subsequent framework-evaluation phase; one clinician contributed to both phases. The co-design discussions focused on the study procedures and materials rather than on rating or prioritising the final outcome domains.

Contributor feedback resulted in several modifications before the main evaluation phase. Regarding outcome-measure selection, contributors considered the five identified PROMs to contain relevant and complementary concepts but raised concerns about the burden and practicality of administering multiple complete questionnaires. The five measures were therefore retained as source instruments for the structured synthesis, with selected clinically and conceptually relevant content taken forward for mapping rather than administering the complete instruments to contributors.

The co-design process also identified areas requiring greater emphasis within the preliminary framework. These included prosthesis usability, particularly donning and doffing, and the influence of environmental and contextual factors on mobility, including terrain, weather, and social context. Terminology was refined to improve accessibility, including adoption of the term “prosthesis experience” and clearer plain-language descriptions of mobility and psychosocial wellbeing.

Feedback further informed the domain-rating exercise. Contributors considered the distinction between importance and actionability useful but identified a need to define actionability more explicitly. Accordingly, actionability was operationalised as the perceived practicality of assessing or monitoring a domain within regular care, with plain-language explanations and examples incorporated into the main evaluation materials. Recruitment procedures were also broadened to include community-based and user-led routes, and provision for language support was incorporated to facilitate participation where required. Collectively, these modifications shaped the framework, materials, and procedures subsequently used in the main study.

### 3.2 Step 1: Selected PROM Content Synthesis and Framework Development

The structured synthesis comprised 112 purposively selected content units drawn from the PEQ, TAPES, PLUS-M, OPUS, and LCI-5. These units represented clinically and conceptually relevant content selected to inform development of the preliminary framework and should not be interpreted as an exhaustive item-level assessment of the complete instruments.

Across the selected content, 42 units (37.5%) were assigned to Mobility, 25 (22.3%) to Physical Function, 19 (17.0%) to Psychosocial Wellbeing, and 26 (23.2%) to Prosthesis Experience. The distribution differed substantially between source instruments (Figure 1). Within the selected PEQ content, all four domains were represented, with the greatest number of mapped units relating to Prosthesis Experience (12) and Mobility (10). Selected TAPES content placed greater emphasis on Psychosocial Wellbeing (10 of 23 mapped units), while PLUS-M content was predominantly assigned to Mobility (13 of 18). OPUS showed comparatively broad representation of Physical Function (11 of 27) and Prosthesis Experience (9 of 27), whereas the selected LCI-5 content was confined to Mobility (8 of 12) and Physical Function (4 of 12).

**Figure 1:**
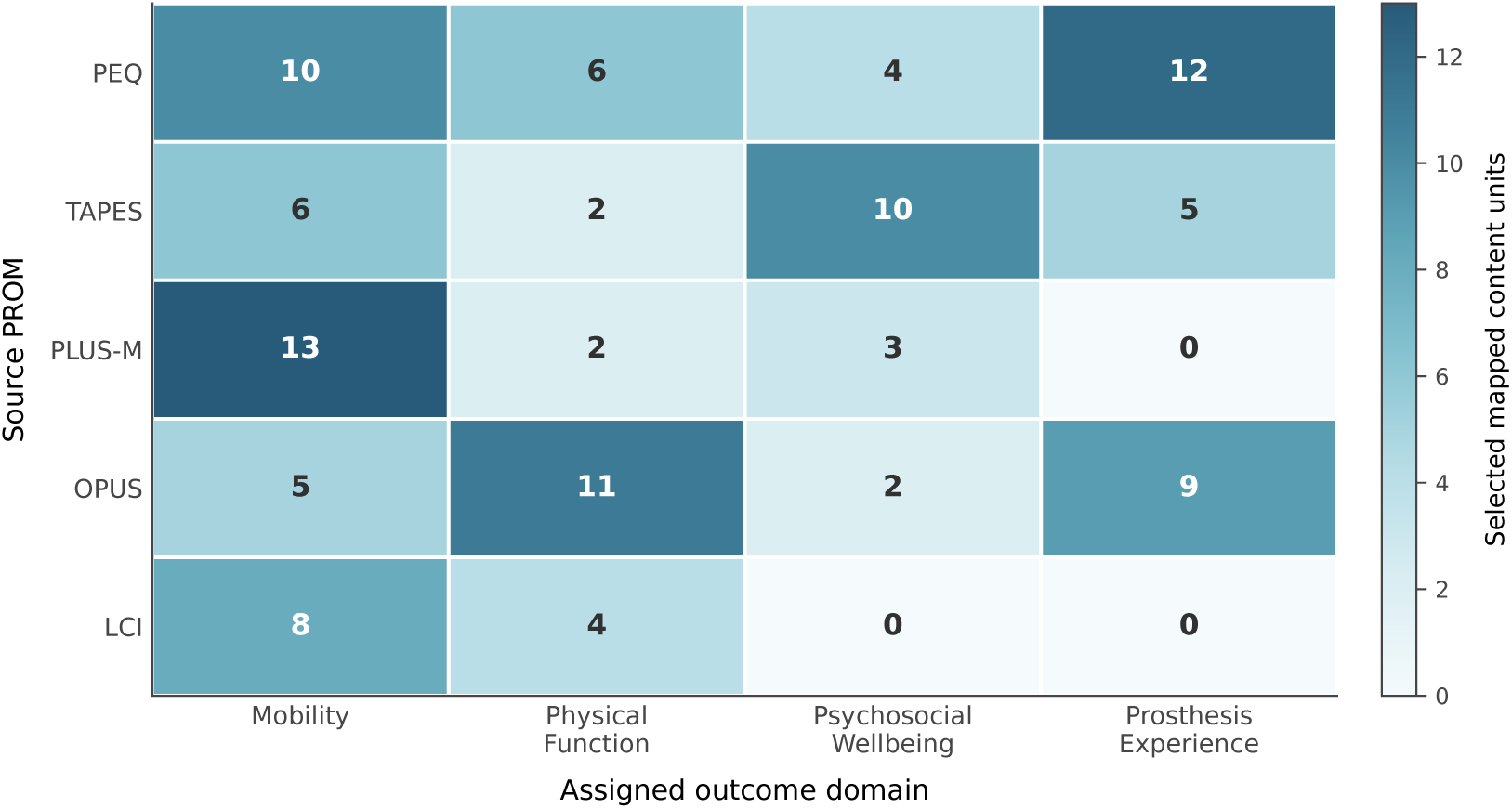
Distribution of purposively selected PROM content across the four outcome domains. Cell values indicate the number of selected content units mapped to each domain; counts should not be interpreted as exhaustive coverage of the complete instruments.

At the theme and subtheme level, the selected Mobility content most frequently represented terrain negotiation (*n* = 9), walking capacity (*n* = 8), stairs (*n* = 6), endurance (*n* = 5), and outdoor mobility (*n* = 5). Within Physical Function, transfers (*n* = 7) and personal care (*n* = 5) were most frequently represented. Social participation (*n* = 6) was the most frequent Psychosocial Wellbeing theme, while confidence, emotional adjustment, self-image, fear of falling, identity, mental health, and social support were represented to varying degrees. Within Prosthesis Experience, the most frequent themes were socket fit (*n* = 4), usability (*n* = 4), aesthetics (*n* = 3), comfort (*n* = 3), and overall satisfaction (*n* = 3).

The synthesis therefore demonstrated substantial variation in the types of concepts represented within the selected content from different PROMs. Mobility-related concepts were prominent across several measures, whereas psychosocial and prosthesis-related concepts were concentrated more strongly within particular instruments. Coding and grouping of the selected content resulted in the four-domain preliminary framework comprising Mobility, Physical Function, Psychosocial Wellbeing, and Prosthesis Experience, with associated themes and illustrative examples as presented in Table 2. This framework, incorporating the terminology and procedural refinements arising from the preparatory co-design phase, was subsequently taken forward for stakeholder evaluation in Step 2.

### 3.3 Step 2: Stakeholder Interviews and Domain Evaluation

#### 3.3.1 Overview of Contributors

A total of 18 contributors participated in the main framework-evaluation phase, comprising twelve prosthesis users (U1–U12), four clinicians (C1–C4), and two industrial partners (IP01–IP02). These contributors evaluated the preliminary four-domain framework through semi-structured interviews and structured ratings of domain importance and actionability. The prosthesis-user group ranged in age from 29 to 73 years and included individuals with differing levels and causes of lower-limb loss or absence and between two and fourteen years of prosthesis use. Participants included individuals with transtibial and transfemoral limb loss, one bilateral transtibial user, and one participant with congenital limb absence. Causes of acquired limb loss included trauma, vascular disease, diabetes, infection, and cancer. This variation enabled perspectives to be obtained across differing durations and experiences of prosthesis use.

Mobility support also varied substantially. Three contributors reported no regular mobility aid, while others used sticks, crutches, wheelchairs, or combinations of these depending on the activity and environment. Some aids were used only occasionally or for longer outdoor distances. The sample also reflected varied ethnic, socioeconomic, and living circumstances, including contributors from refugee and asylum-seeking backgrounds. To reduce the risk of re-identification when participant identifiers are used alongside quotations, detailed ethnicity and living circumstances are reported only at group level rather than linked to individual identifiers. Table 3 summarises the principal clinical characteristics of the prosthesis-user group.

**Table 3:**
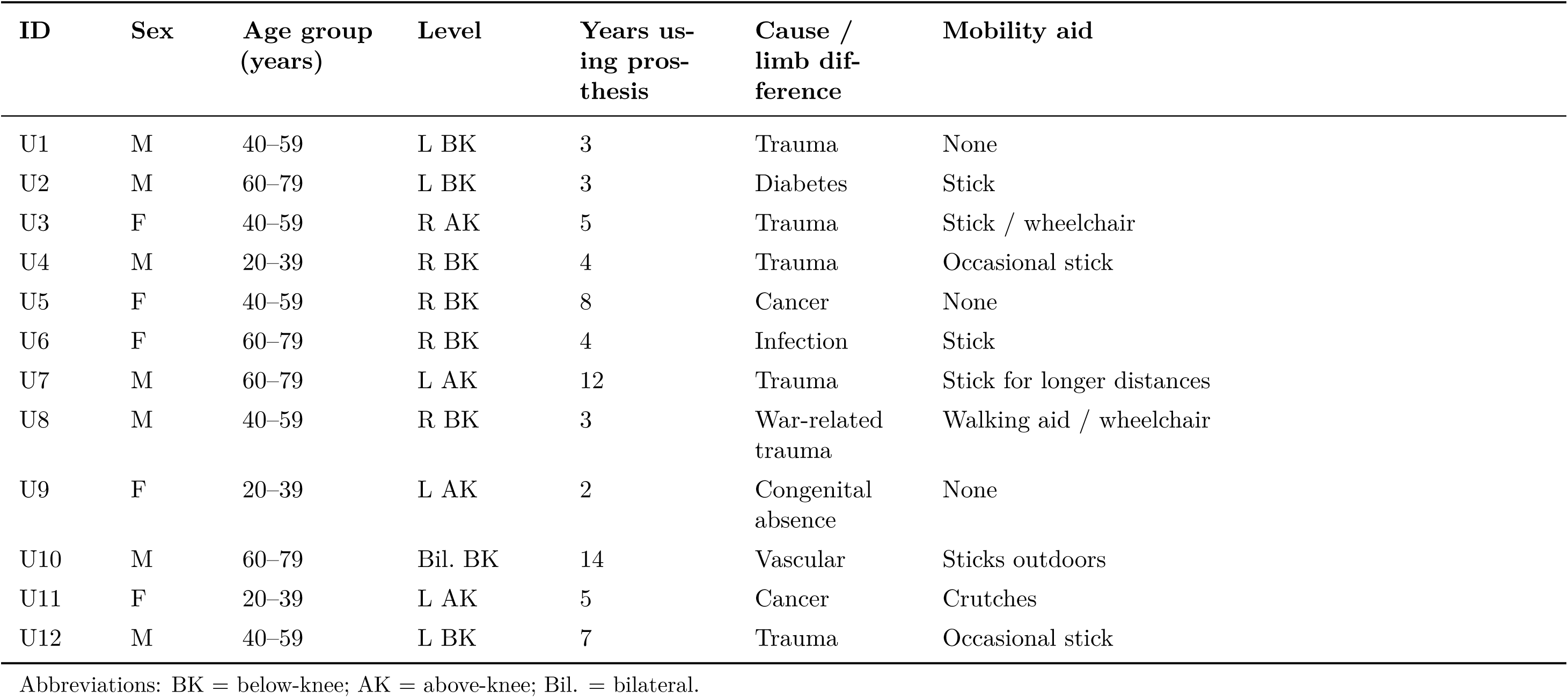
Clinical characteristics of prosthesis-user contributors in the main evaluation phase (*n* = 12).

Four clinicians participated in the main evaluation phase, representing expertise across prosthetics, physiotherapy, rehabilitation medicine, and clinically relevant rehabilitation research. Their professional experience ranged from eight to twenty years and included direct clinical practice, teaching-hospital activity, and research-based roles. The group comprised a prosthetist (C1), a physiotherapist (C2), a rehabilitation consultant (C3), and a clinical researcher working within a university-linked teaching-hospital setting (C4). Table 4 summarises their professional backgrounds.

**Table 4:** Clinician contributors in the main evaluation phase (*n* = 4).

| ID | Role | Experience<br>(years) | Setting | Outcome measures/tools used | Real-world<br>monitoring |
| --- | --- | --- | --- | --- | --- |
| C1 | Prosthetist | 10–15 | Rehabilitation centre | Comfort assessment; VAS | No |
| C2 | Physiotherapist | 8 | Hospital outpatient rehabilitation | AMP Pro; TUG | Occasionally |
| C3 | Rehabilitation consultant | 12 | Community rehabilitation | VAS; GAS | No |
| C4 | Clinical researcher | 20 | University-linked teaching hospital and rehabilitation research | AMP Pro; PROMs | No |

VAS = Visual Analogue Scale; AMP Pro = Amputee Mobility Predictor; TUG = Timed Up and Go; GAS = Goal Attainment Scaling; PROMs = patient-reported outcome measures.

Two industrial partners participated in the main evaluation phase, providing complementary perspectives from prosthetic device design and commercial product development. Their experience ranged from four to seven years and included both technical and market-facing interaction with prosthesis users. One contributor held a technical and consultancy role in prosthetic design, while the other worked in product and sales development. Their professional profiles are summarised in Table 5.

**Table 5:** Industrial partner contributors in the main evaluation phase (*n* = 2).

| **ID** | **Professional role** | **Primary area of expertise** | **Experience (years)** |
| --- | --- | --- | --- |
| IP01 | Technical and consultancy role | Prosthetic design and development | 7 |
| IP02 | Product and sales role | Prosthetic products and user-facing development | 4 |

#### 3.3.2 Qualitative Interview Findings

Across the interviews, contributors described the four domains as distinct but closely interconnected aspects of everyday prosthesis use and rehabilitation. Table 6 summarises the principal subthemes, influencing factors, and perspectives identified across prosthesis users, clinicians, and industrial partners.

**Table 6:**
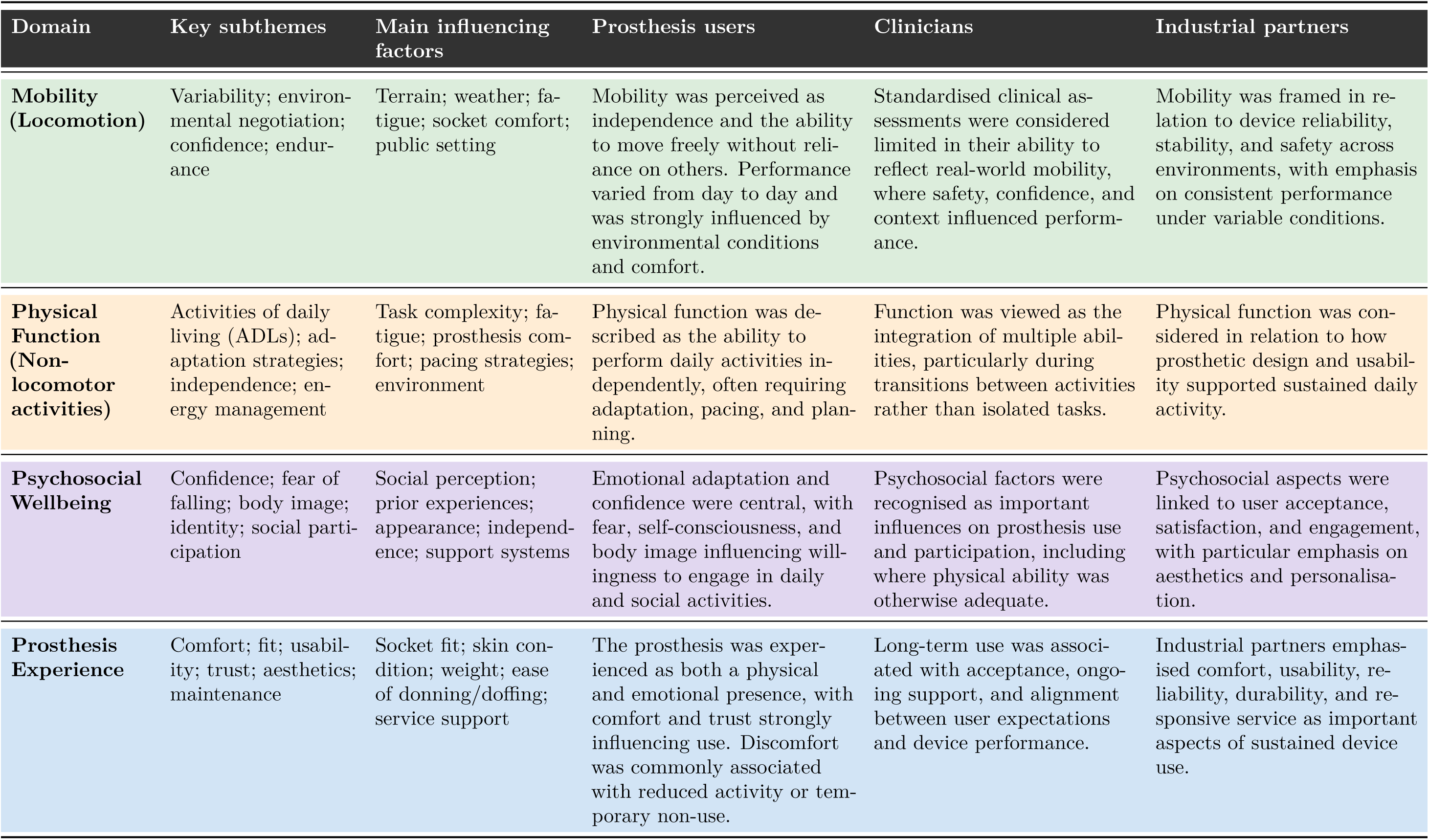
Summary of qualitative interview findings across the four outcome domains.

### Mobility

Mobility was consistently described by contributors as a marker of autonomy in everyday life, extending beyond discrete walking tasks to encompass the ability to move freely and spontaneously within familiar and unfamiliar environments. For most prosthesis users, the significance of mobility lay not in physical performance alone, but in the freedom it conferred, particularly the capacity to leave the home and engage in routine activities without reliance on other people. Several contributors (U1, U3, U4, U6) identified independent community access as a meaningful indicator of rehabilitation progress. “Walking is one thing, but it is more about being able to just go when I want.” (U3). “When I can go to the shop or meet a friend without asking for help, that feels like real progress.” (U4).

Mobility extended beyond routine community activities such as shopping and collecting medication to include work and caring responsibilities, use of public transport, and attendance at appointments. Walking ability was described as fluctuating considerably from day to day, shaped by socket comfort, fatigue, pain, and environmental conditions. Several contributors (U2, U5, U7, U9) described this variability as a persistent feature of everyday prosthesis use rather than an exception. “Some days the leg feels alright and I walk fine, then the next day it rubs and I hardly move.” (U5). “It really depends how my stump feels. If it is sore, I do less and try again later.” (U9). Some contributors also described occasions on which poor mobility days affected their ability to attend rehabilitation or prosthetic review appointments.

Endurance was frequently discussed, although contributors often described it in relation to both physical effort and the attentional demands of walking. Sustained walking could require continuous attention to balance, foot placement, and prosthesis comfort. “You get tired from thinking about every step, not just the walking.” (U6). “I try to go as far as I can, but after a bit the pressure builds and I know I should stop.” (U8).

Environmental conditions were repeatedly identified as important influences on mobility. Uneven surfaces, slopes, and wet conditions were described as physically and psychologically demanding, while public spaces introduced additional concerns related to fear of falling and social visibility. “I do not like walking on grass or stones. You have to watch where you are putting your feet all the time.” (U7). “On pavements I am fine, but if it is a hill or gravel, I slow right down.” (U10).

Social context further shaped mobility experience. Several contributors described heightened self-consciousness when walking in public and reported modifying their movement in response to perceived observation by others. In some instances, these adaptations were associated with increased discomfort or fatigue. “You try to look steady because people stare if you wobble.” (U3). “Sometimes I walk faster just to look alright, then I get sore after.” (U8).

Clinicians similarly described real-world mobility as both measurable and contextually complex. Several clinicians (C1, C2, C3) considered performance observed in controlled or clinical settings to provide an incomplete representation of how individuals moved in everyday environments. “We can measure walking speed, but that does not show how safe someone feels in real conditions.” (C2). “In the lab they look steady, but outside there are slopes, distractions, and people watching. It changes everything.” (C3). Clinicians also described gradual changes in mobility that could be difficult to identify without regular follow-up, including increased caution, altered walking strategies, and avoidance of more challenging environments. “You see them six months later and they are slower, more cautious. Without follow-up, you would not realise it happened.” (C1).

Industrial partners emphasised safety, stability, and reliability across variable environmental conditions. Their accounts focused particularly on whether devices performed consistently across uneven terrain, slopes, and changing weather conditions, and on the importance of predictable device behaviour in supporting user confidence and continued mobility.

### Physical Function

Physical function was consistently described as the ability to manage everyday life independently, encompassing non-locomotor activities that structured contributors’ daily routines. Tasks such as cooking, cleaning, gardening, and household maintenance were identified as meaningful indicators of progress, with independent performance associated with a sense of normality and self-sufficiency. “It is not really about walking far; it is about getting through the day and still having some energy left.” (U9). “When I can do simple things like make a meal or clean up without asking for help, that feels like I am doing okay.” (U3).

Many contributors reported that completing everyday activities required adaptation, pacing, and planning. “I can do most things but slower. I just take my time now.” (U5). “I do one thing, sit down for a bit, then carry on. It is the only way to manage the day.” (U6). Prosthesis comfort also influenced engagement in daily activities, with contributors describing differences in whether the device was required for particular tasks. Some static or seated activities could be completed without the prosthesis, whereas standing tasks and movement around the home more commonly required its use, indicating variation in prosthesis dependence across everyday activities.

Functional ability was also linked to self-image and a sense of purpose. Contributors associated domestic contribution, hobbies, and the ability to assist others with regaining valued aspects of their pre-existing roles. “Getting back to doing small things, like fixing stuff or helping out, makes me feel useful.” (U1). “Even if it is just pottering around in the garden, it gives me a bit of pride.” (U9).

Clinicians conceptualised physical function as a multidimensional construct involving coordination, balance, endurance, and the ability to transition between activities. They emphasised that everyday function rarely occurred as a sequence of isolated tasks. “We test walking or standing, but life does not happen one task at a time.” (C4). “You see them perform great in the clinic, then you find out they avoid things at home because it is tiring or awkward.” (C2).

Industrial partners similarly emphasised usability, including the practical demands of donning and doffing the prosthesis. Their accounts highlighted the importance of device design in supporting routine activity and accommodating variation between activities for which the prosthesis was essential and those that could be undertaken without it.

### Psychosocial Wellbeing

Psychosocial wellbeing was consistently described, particularly by prosthesis users, as central to rehabilitation and everyday prosthesis use. Recovery was characterised not only as a physical process but also as an ongoing adjustment involving identity, confidence, self-image, and social participation. Contributors described frustration, reduced confidence, and heightened awareness of being observed in public. “You can have the best leg, but if you do not feel right about it, you will not use it properly.” (U5). “Sometimes it is not the pain; it is just the feeling that people are looking at you.” (U3).

Confidence was described as dynamic and sensitive to setbacks, influencing willingness to engage in everyday and social activities. Falls or episodes of discomfort could undermine confidence that had taken considerable time to rebuild. “You have a good run for a few weeks, then one fall and it sets you back.” (U4). “I think I am doing fine until something goes wrong, and then I feel like I am starting over.” (U6).

The psychosocial consequences of prosthesis use were also evident in work, parenting, and other valued social roles. Contributors described adapting their expectations and routines while continuing to identify strongly with these roles. “My confidence drops around hour six. There is always a low-level fear that a stumble will turn into a fall in front of everyone. But I have accepted that doing the job at a slower pace is better than not doing it at all.” (U4). “My identity as an active parent is still there, but it looks different now, more planning, less spontaneity. Over time, I have accepted that careful is not the same as incapable.” (U11).

Body image was a recurring theme, with contributors describing how prosthesis appearance influenced confidence and willingness to participate socially. “At the beginning, I always wore long trousers to hide it. Now I just wear what I want.” (U9). “If the leg looks bulky, I do not like going out much. I prefer when it looks neat.” (U7). Regaining independence was similarly described as an important psychosocial milestone. “The first time I drove again, I felt like me, not a patient.” (U2). “Doing things on my own again makes me feel normal, even if it is small stuff.” (U8).

Clinicians described emotional factors, including fear of falling and embarrassment, as capable of restricting participation even where physical ability appeared adequate. They also emphasised expectation management during rehabilitation. “Confidence makes the difference between being able to walk and actually doing it.” (C3). “We try to explain early on that it takes time to feel natural. That prevents frustration later.” (C4).

Industrial partners discussed psychosocial wellbeing primarily through device acceptance, aesthetics, and personalisation. Their accounts suggested that the appearance and perceived individuality of a prosthesis could influence how comfortably users incorporated the device into social and everyday life.

### Prosthesis Experience

Prosthesis experience was described as both a physical and emotional process, centred on comfort, trust, usability, and adaptation over time. Contributors emphasised that successful use depended not only on technical fit but also on developing familiarity and confidence with the device. “At first it felt strange, like it was not really mine. Now I am used to it, but some days it still feels awkward.” (U6). “When it fits right and works how it should, I do not think about it anymore.” (U4).

Comfort was among the most consistently discussed influences on prosthesis use. Discomfort, soreness, heat, sweating, and skin irritation could lead contributors to remove the prosthesis earlier than intended or reduce their activity for the remainder of the day. “If it starts rubbing, I just take it off. It ruins the rest of the day.” (U5). “When it feels good, I walk more. When it does not, I sit more.” (U9).

Trust in the device was similarly important. Negative experiences could affect confidence in subsequent use, particularly where contributors were uncertain about stability or reliability. “You have to be able to trust it; one bad step and it is hard to get that trust back.” (U8).

Appearance influenced prosthesis experience in ways that extended beyond cosmetic preference. Contributors described the visual presentation of the device as affecting self-consciousness, confidence, and willingness to participate in public settings. Personalisation could support a greater sense of acceptance and ownership. “I used to cover it up, but now I got one that looks nice, so I do not care.” (U3). “Making it look more personal helps. It feels less like hospital stuff.” (U7).

Ease of use also shaped daily engagement. Donning, doffing, cleaning, and adjusting the prosthesis formed part of contributors’ everyday routines, and burdensome preparation could reduce motivation to use the device. “If it takes me ages to put on, I just do not bother going out that day.” (U1).

Clinicians highlighted the importance of developing a sense of ownership and acceptance alongside achieving appropriate technical fit. “You can have a perfect fit, but if they do not feel connected to it, they will not wear it.” (C4). “When someone accepts it as part of them, that is when rehabilitation really works.” (C1). Follow-up care was also described as important for maintaining engagement during periods of discomfort or adjustment. “Good follow-up keeps people engaged. If they feel ignored, they stop using it.” (C3).

Industrial partners framed prosthesis experience around usability, reliability, comfort, aesthetics, and user-centred design. Their accounts emphasised the importance of integrating the device into the user’s everyday routines rather than considering technical performance in isolation. As one industrial partner described, long-term engagement depended on whether the device “fits seamlessly into the user’s daily routine and expectations” (IP02).

#### 3.3.3 Cross-Domain Interdependence

Across interviews, Mobility, Physical Function, Psychosocial Wellbeing, and Prosthesis Experience were described as closely and dynamically interconnected. Contributors consistently reported that changes in one domain could influence experience in others. Improvements in comfort or mobility were commonly associated with greater confidence and engagement in everyday activity, whereas pain, fatigue, discomfort, or reduced confidence could restrict participation. “When I walk better, everything feels better. It is like a boost.” (U5). “If the leg hurts, I stop using it, then I lose confidence again.” (U4).

Importantly, positive outcomes in one domain did not necessarily correspond to positive outcomes across all others. One prosthesis user described having adequate mobility in some environments while experiencing substantial difficulties maintaining and using the prosthesis within their living environment. The contributor explained: “The leg works fine on the road. But here, the problem is not walking. The problem is sleeping, washing, and not losing it when someone steps on it in the dark.” (prosthesis user). This account illustrated how environmental circumstances and prosthesis-management demands could constrain everyday use despite adequate walking ability.

Clinicians similarly described situations in which apparently strong mobility performance coexisted with unmet psychosocial or prosthesis-related needs. One clinician recalled an individual who performed well on clinical mobility assessments but repeatedly avoided rehabilitation because of distress related to the appearance of the prosthesis. “I almost discharged her last month because her walking was fine. But she was hiding a whole different problem. Our job is not just metres walked. It is whether they walk where they want to be seen.” (C4). Such accounts demonstrated that performance within one outcome domain could provide an incomplete representation of the individual’s broader rehabilitation experience.

Clinicians characterised rehabilitation as a dynamic interaction between physical, psychosocial, and prosthesis-related factors. “Comfort, confidence, and movement are all tied together. You cannot fix one without looking at the rest.” (C1). “People might walk well in clinic but avoid stairs or outdoor surfaces because of fear or fatigue.” (C3).

Industrial partners described a similar interdependence from a design perspective, emphasising relationships between device performance, usability, aesthetics, and user perception. Across stakeholder groups, successful adaptation was therefore characterised less as optimisation of a single outcome and more as the ability to sustain everyday prosthesis use across changing physical, psychosocial, environmental, and device-related circumstances.

#### 3.3.4 Comparative Perspectives Across Stakeholder Groups

Comparison across prosthesis users, clinicians, and industrial partners showed substantial agreement on the relevance of all four domains, but differences in how each stakeholder group framed the outcomes that mattered. These differences reflected stakeholder roles and experiences rather than disagreement about the overall importance of the framework. Mobility was a shared priority across all three groups. Prosthesis users primarily described mobility in terms of independence, spontaneity, confidence, and the ability to participate in community life. Clinicians focused more strongly on the discrepancy between performance observed in clinical settings and mobility experienced under real-world environmental conditions, including the difficulty of detecting gradual functional change without regular follow-up. Industrial partners approached mobility principally through device stability, reliability, and predictable performance across variable environments.

Physical Function was similarly valued across stakeholder groups, although the emphasis differed. Prosthesis users described successful function through the ability to complete everyday activities independently while managing fatigue, pacing, and adaptation. Clinicians emphasised the multidimensional and sequential nature of everyday function, particularly the limitations of assessing isolated tasks when daily life requires transitions between activities. Industrial partners focused more strongly on usability and the extent to which device design, including ease of donning and doffing, supported sustained participation in routine activities.

Psychosocial Wellbeing was particularly prominent in the accounts of prosthesis users and clinicians. Users described confidence, body image, identity, fear of falling, social visibility, and participation as integral components of everyday prosthesis use. Clinicians similarly recognised that fear, embarrassment, confidence, and expectations could influence participation even when physical capability appeared adequate. Industrial partners discussed psychosocial outcomes less directly, tending instead to frame them through device acceptance, aesthetics, personalisation, and the relationship between product design and willingness to use the prosthesis.

Prosthesis Experience was strongly represented across all three stakeholder groups but was again framed differently. Prosthesis users emphasised comfort, trust, fit, ease of use, maintenance, and the gradual development of familiarity with the device. Clinicians focused on the interaction between appropriate technical fit, user acceptance, ownership, and continuing follow-up. Industrial partners placed particular emphasis on comfort, reliability, usability, durability, aesthetics, and the integration of the prosthesis into everyday routines.

Taken together, the stakeholder comparison indicated broad convergence around independence, confidence, comfort, safety, and participation, alongside complementary stakeholder-specific perspectives. Prosthesis users contributed detailed accounts of lived experience and day-to-day variability; clinicians highlighted assessment, follow-up, and the relationship between clinical performance and everyday function; and industrial partners emphasised device performance, usability, reliability, and user-centred design. Table 7 summarises the principal barriers and enabling factors identified across the three groups.

**Table 7:** Comparative barriers and enabling factors identified across prosthesis users, clinicians, and industrial partners.

| Domain | Prosthesis users | Clinicians | Industrial partners | Enabling factors |
| --- | --- | --- | --- | --- |
| **Mobility (Locomotion)** | Pain; fatigue; uneven terrain; fear of falling; social visibility; day-to-day variability | Environmental complexity; reduced confidence; discrepancy between clinical and real-world performance; limited opportunity to observe gradual change | Stability and reliability across terrains; predictable performance under variable environmental conditions | Regular follow-up; confidence in device performance; adaptation to environmental demands; context-sensitive assessment |
| **Physical Function (Non-locomotor activities)** | Fatigue; task complexity; socket discomfort; need for pacing and adaptation | Difficulty representing sequential everyday activities using isolated clinical tasks; balance and endurance demands | Usability limitations; effort associated with donning, doffing, and routine device interaction | Pacing and adaptation strategies; task-relevant rehabilitation; usable device design; support for everyday routines |
| **Psychosocial Wellbeing** | Reduced confidence; fear after falls; self-consciousness; body image concerns; social visibility | Fear of falling; embarrassment; reduced confidence; unrealistic or unmet expectations | Aesthetic and personalisation considerations; relationship between device design and acceptance | Expectation management; confidence-building; social support; personalisation; restoration of valued roles |
| **Prosthesis Experience** | Discomfort; poor fit; skin irritation; reduced trust; maintenance burden; difficult donning and doffing | Technical fit; acceptance and ownership; need for continued support and follow-up | Comfort; reliability; usability; durability; aesthetics; alignment between design and everyday routines | Appropriate fit and comfort; responsive follow-up; reliable and usable design; personalisation; integration into daily life |

#### 3.3.5 Quantitative Evaluation of Domain Importance and Actionability

Structured ratings provided complementary quantitative evidence regarding the perceived importance and actionability of the four outcome domains. Importance reflected the perceived relevance of each domain to meaningful prosthesis use and rehabilitation, whereas actionability reflected the perceived practicality of assessing or monitoring the domain within regular care. Ratings were analysed descriptively for prosthesis users (*n* = 12), clinicians (*n* = 4), and industrial partners (*n* = 2). Given the small stakeholder-group sample sizes, particularly for industrial partners, comparisons between groups were descriptive and no inferential statistical testing was undertaken.

Among prosthesis users, Mobility and Physical Function received the highest median importance ratings, both at 5.0. Psychosocial Wellbeing and Prosthesis Experience also received high importance ratings, with medians of 4.5. Actionability ratings showed greater differentiation between domains. Mobility had the highest median actionability rating at 4.5, followed by Physical Function and Psychosocial Wellbeing at 4.0, while Prosthesis Experience had the lowest median at 3.0. The corresponding median importance-actionability differences were 0.5 for Mobility, 1.0 for Physical Function, 0.5 for Psychosocial Wellbeing, and 1.5 for Prosthesis Experience.

Clinicians similarly rated all four domains as important, although the pattern of perceived actionability differed. Mobility and Psychosocial Wellbeing both received median importance ratings of 5.0, followed by Physical Function at 4.5 and Prosthesis Experience at 4.0. Median actionability ratings were 4.0 for Mobility and Physical Function and

3.0 for both Psychosocial Wellbeing and Prosthesis Experience. The largest importance-actionability difference among clinicians was therefore observed for Psychosocial Wellbeing, with a median difference of 2.0, compared with 1.0 for Mobility, 0.5 for Physical Function, and 1.0 for Prosthesis Experience.

The two industrial partners both rated Mobility and Prosthesis Experience at the maximum importance score of 5, resulting in median importance ratings of 5.0 for both domains. Physical Function had a median importance rating of 4.5, while Psychosocial Wellbeing had a median of 3.5. Median actionability ratings were 4.5 for Mobility and Prosthesis Experience, 3.5 for Physical Function, and 2.5 for Psychosocial Wellbeing. Because this stakeholder group comprised only two contributors, these values are presented descriptively alongside the individual observations and should not be interpreted as estimates of a wider industrial perspective.

Across stakeholder groups, Mobility was consistently rated highly for both importance and actionability. Greater divergence between importance and actionability was evident for other domains, but the pattern differed by stakeholder group. Prosthesis users showed their largest median difference for Prosthesis Experience, clinicians for Psychosocial Wellbeing, and industrial partners showed equivalent median differences of 1.0 for Physical Function and Psychosocial Wellbeing. Figure 2 presents the individual ratings together with the descriptive medians and highlights these within-group importance-actionability differences.

**Table 8:** Descriptive importance and actionability ratings across stakeholder groups.

| Domain | Importance (1–5) | Actionability (1–5) | Median difference |
| --- | --- | --- | --- |
| **Prosthesis users ($n = 12$)** |  |  |  |
| Mobility | 5.0 [5.0–5.0]; 4.83 ± 0.39 | 4.5 [4.0–5.0]; 4.42 ± 0.67 | 0.5 |
| Physical Function | 5.0 [4.75–5.0]; 4.75 ± 0.45 | 4.0 [4.0–5.0]; 4.25 ± 0.75 | 1.0 |
| Psychosocial Wellbeing | 4.5 [4.0–5.0]; 4.42 ± 0.67 | 4.0 [3.0–4.0]; 3.58 ± 0.67 | 0.5 |
| Prosthesis Experience | 4.5 [4.0–5.0]; 4.42 ± 0.67 | 3.0 [3.0–4.0]; 3.42 ± 0.79 | 1.5 |
| **Clinicians ($n = 4$)** |  |  |  |
| Mobility | 5.0 [4.75–5.0]; 4.75 ± 0.50 | 4.0 [3.75–4.25]; 4.00 ± 0.82 | 1.0 |
| Physical Function | 4.5 [4.0–5.0]; 4.50 ± 0.58 | 4.0 [3.75–4.25]; 4.00 ± 0.82 | 0.5 |
| Psychosocial Wellbeing | 5.0 [4.75–5.0]; 4.75 ± 0.50 | 3.0 [2.75–3.25]; 3.00 ± 0.82 | 2.0 |
| Prosthesis Experience | 4.0 [3.75–4.25]; 4.00 ± 0.82 | 3.0 [2.75–3.25]; 3.00 ± 0.82 | 1.0 |
| **Industrial partners ($n = 2$)** |  |  |  |
| Mobility | 5.0 [5–5] | 4.5 [4–5] | 0.5 |
| Physical Function | 4.5 [4–5] | 3.5 [3–4] | 1.0 |
| Psychosocial Wellbeing | 3.5 [3–4] | 2.5 [2–3] | 1.0 |
| Prosthesis Experience | 5.0 [5–5] | 4.5 [4–5] | 0.5 |

For prosthesis users and clinicians, values are presented as median [IQR]; mean ± SD. For industrial partners, median [range] is reported because only two contributors were included. Median differences were calculated as importance minus actionability and are descriptive rather than inferential.

**Figure 2:**
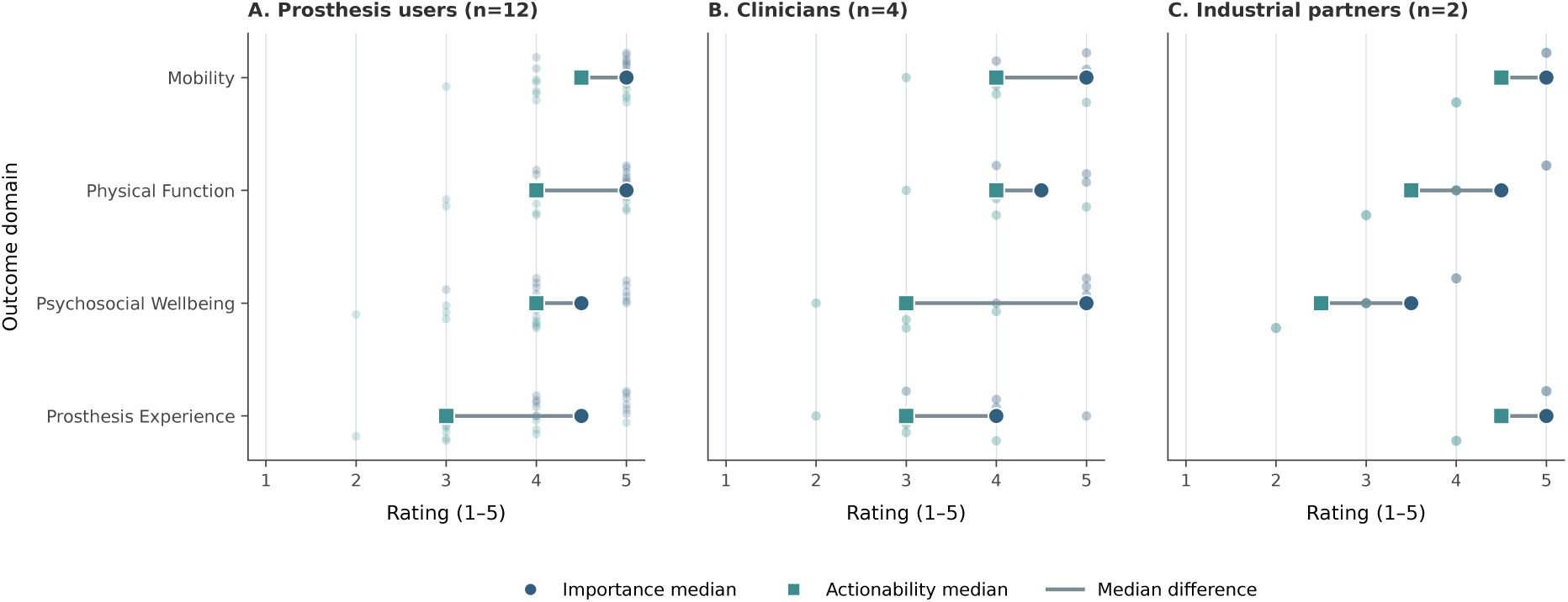
Importance and actionability ratings across stakeholder groups. Large circles and squares indicate median importance and actionability ratings, respectively; smaller points show individual ratings and connecting lines indicate median differences. Industrial-partner findings (*n* = 2) are descriptive.

The distribution of prosthesis-user responses provided further detail regarding the pattern underlying the summary statistics (Figure 3). Ten of twelve users (83.3%) assigned Mobility the maximum importance rating of 5, while nine (75.0%) did so for Physical Function. Psychosocial Wellbeing and Prosthesis Experience each received an importance rating of 4 or 5 from eleven of twelve users (91.7%), demonstrating that their slightly lower medians did not reflect low perceived relevance.

**Figure 3:**
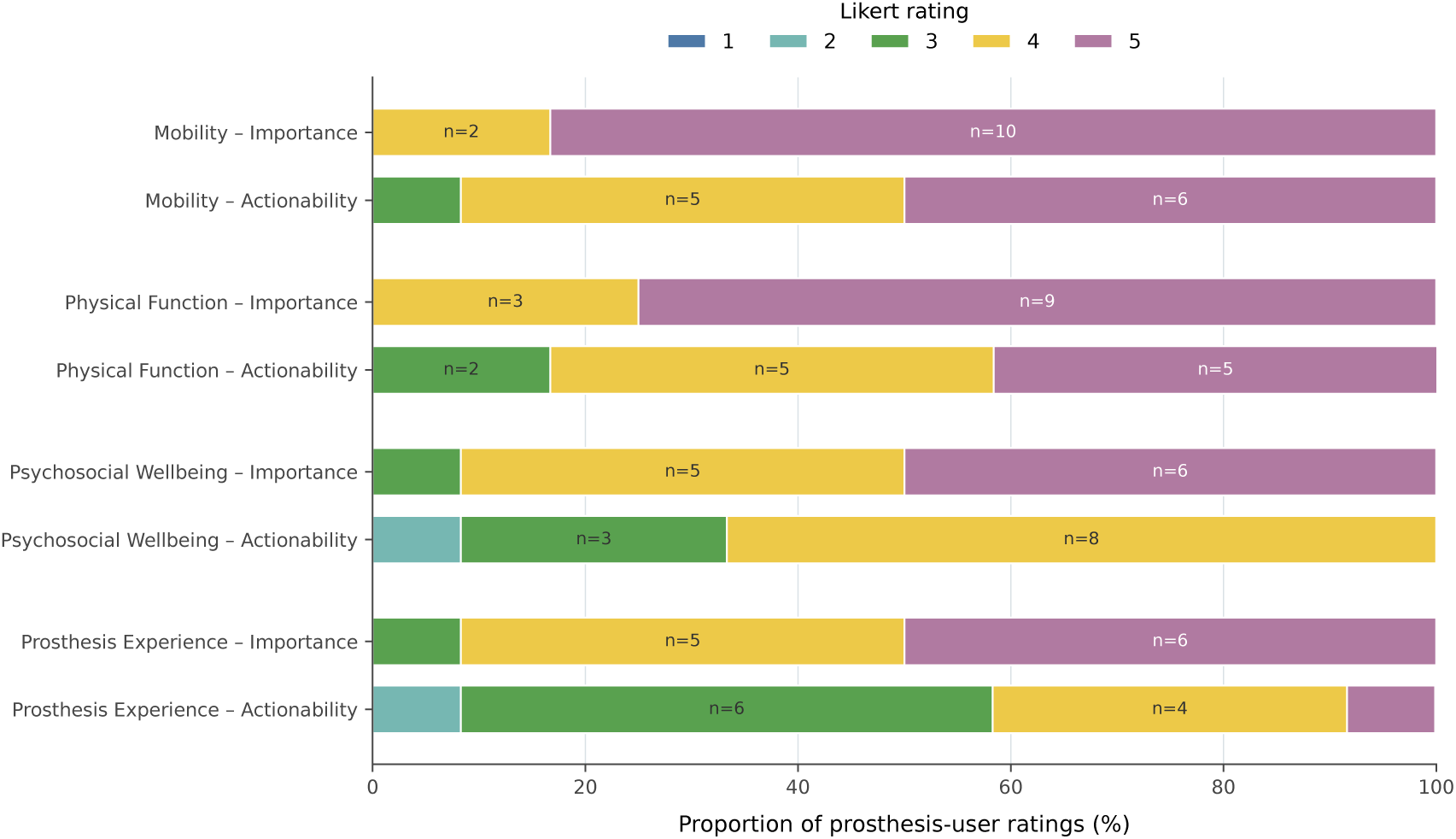
Distribution of prosthesis-user importance and actionability ratings across the four outcome domains (*n* = 12).

Greater dispersion was apparent in the actionability ratings. For Mobility, eleven of twelve users (91.7%) selected ratings of 4 or 5, compared with ten of twelve (83.3%) for Physical Function. Psychosocial Wellbeing actionability was concentrated at rating 4, selected by eight users (66.7%), with no user assigning the maximum rating of 5. Prosthesis Experience showed the broadest shift towards lower actionability: six users (50.0%) selected rating 3, four (33.3%) selected rating 4, one (8.3%) selected rating 2, and only one (8.3%) selected rating 5. This distribution was consistent with Prosthesis Experience showing the largest median importance-actionability difference within the user group.

### 3.4 Step 3: Integration of Findings

Integration of the selected PROM-content synthesis, qualitative interviews, and quantitative ratings supported retention of all four domains in the final framework while also identifying differences in how strongly particular concepts were represented, experienced, and considered actionable. Across the three strands, Mobility showed the clearest convergence: it accounted for the largest proportion of the selected PROM content, was consistently described as fundamental to independence and community participation, and received high importance and actionability ratings across all three stakeholder groups. Physical Function was similarly supported across strands, particularly through concepts relating to transfers, personal care, everyday activity, fatigue, pacing, and adaptation.

Psychosocial Wellbeing showed a different pattern. Psychosocial concepts appeared less frequently within the selected PROM content than Mobility, and were concentrated more strongly within particular source instruments. In contrast, interviews identified confidence, identity, body image, fear of falling, social visibility, and participation as prominent influences on everyday prosthesis use. Quantitative ratings reinforced their perceived relevance, particularly among prosthesis users and clinicians. Clinicians assigned Psychosocial Wellbeing a median importance rating of 5.0 but a median actionability rating of 3.0, representing the largest importance-actionability difference observed within that stakeholder group. This pattern suggested a distinction between recognising psychosocial outcomes as important and perceiving them as straightforward to assess or monitor within regular care.

Prosthesis Experience was also strongly supported by the integrated findings. Selected content relating to this domain was concentrated particularly within the PEQ and OPUS and included fit, comfort, usability, aesthetics, and satisfaction. Interviews expanded these concepts to include trust, maintenance, ease of donning and doffing, familiarity with the device, service support, and the emotional process of incorporating the prosthesis into everyday life. Prosthesis users showed their largest median importance-actionability difference for this domain, with median ratings of 4.5 for importance and 3.0 for actionability. In contrast, the two industrial partners rated Prosthesis Experience highly for both importance and actionability, reflecting their stronger emphasis on device design, usability, reliability, and product-related factors.

Table 9 summarises the convergence and complementary findings across the three analytic strands. The integrated analysis indicated that the four domains were not experienced as independent outcomes. Mobility and Physical Function were influenced by comfort, confidence, fatigue, environmental demands, and device reliability, while Psychosocial Wellbeing and Prosthesis Experience could affect whether physical capability translated into meaningful everyday participation. Internal and external contextual influences therefore operated across the four domains rather than forming separate outcomes themselves.

**Table 9:**
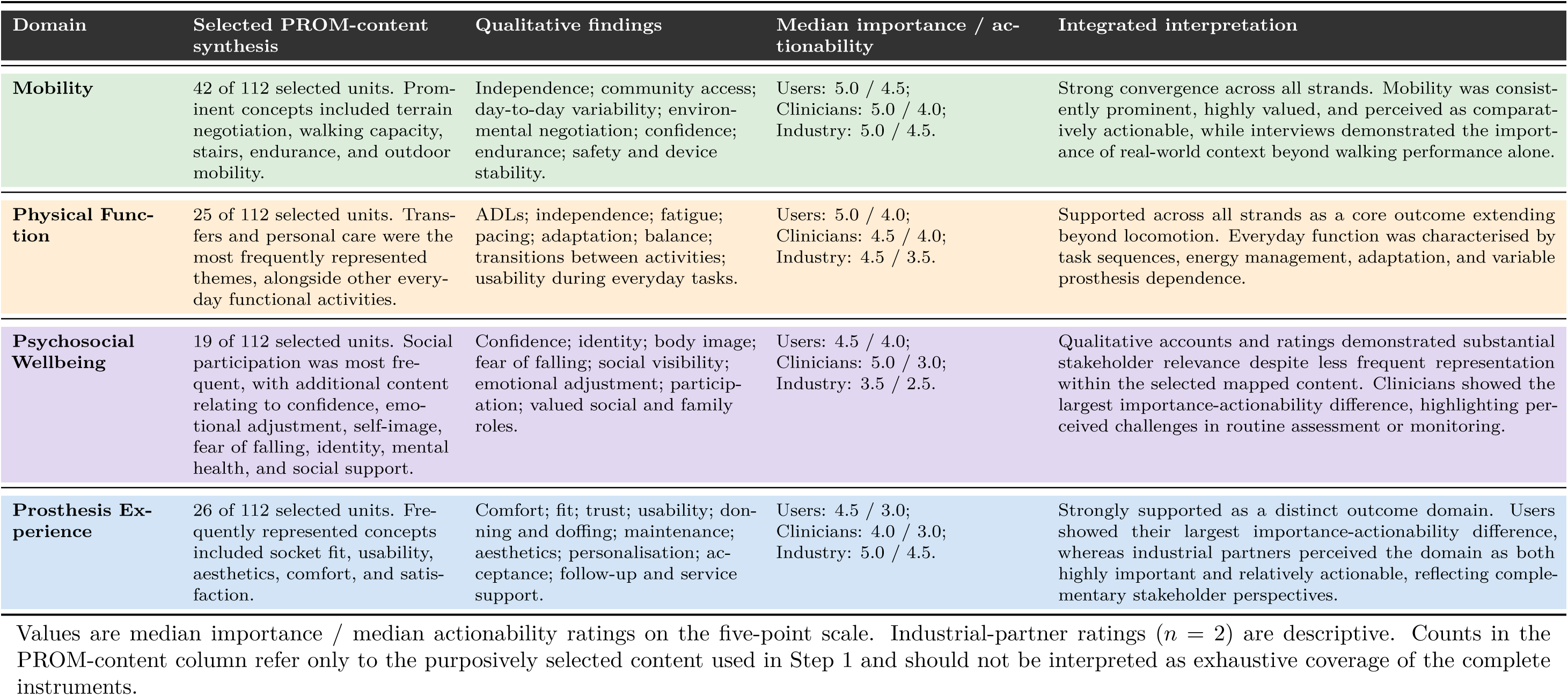
Integration of selected PROM content, qualitative findings, and stakeholder ratings across the four outcome domains.

The integrated findings therefore supported a context-aware four-domain framework in which Mobility, Physical Function, Psychosocial Wellbeing, and Prosthesis Experience represent complementary but interacting dimensions of meaningful everyday prosthesis use. The framework retains the four domains as conceptually distinct to support assessment and interpretation, while recognising that changes within one domain may influence outcomes in others. Contextual influences, including pain, fatigue, environmental conditions, social circumstances, access to support, and characteristics of the prosthesis itself, may modify these relationships across everyday situations.

Figure 4 presents the integrated summary figure derived from the selected PROM-content synthesis, qualitative findings, and structured stakeholder ratings. The four outcome domains are shown as distinct but interrelated components of meaningful everyday prosthesis use. The bidirectional arrows indicate cross-domain interdependence identified in the interviews, while the quantitative annotations highlight selected results that distinguished each domain in the integrated analysis, including the proportion of selected PROM content represented within each domain and the most notable importance–actionability gaps.

**Figure 4:**
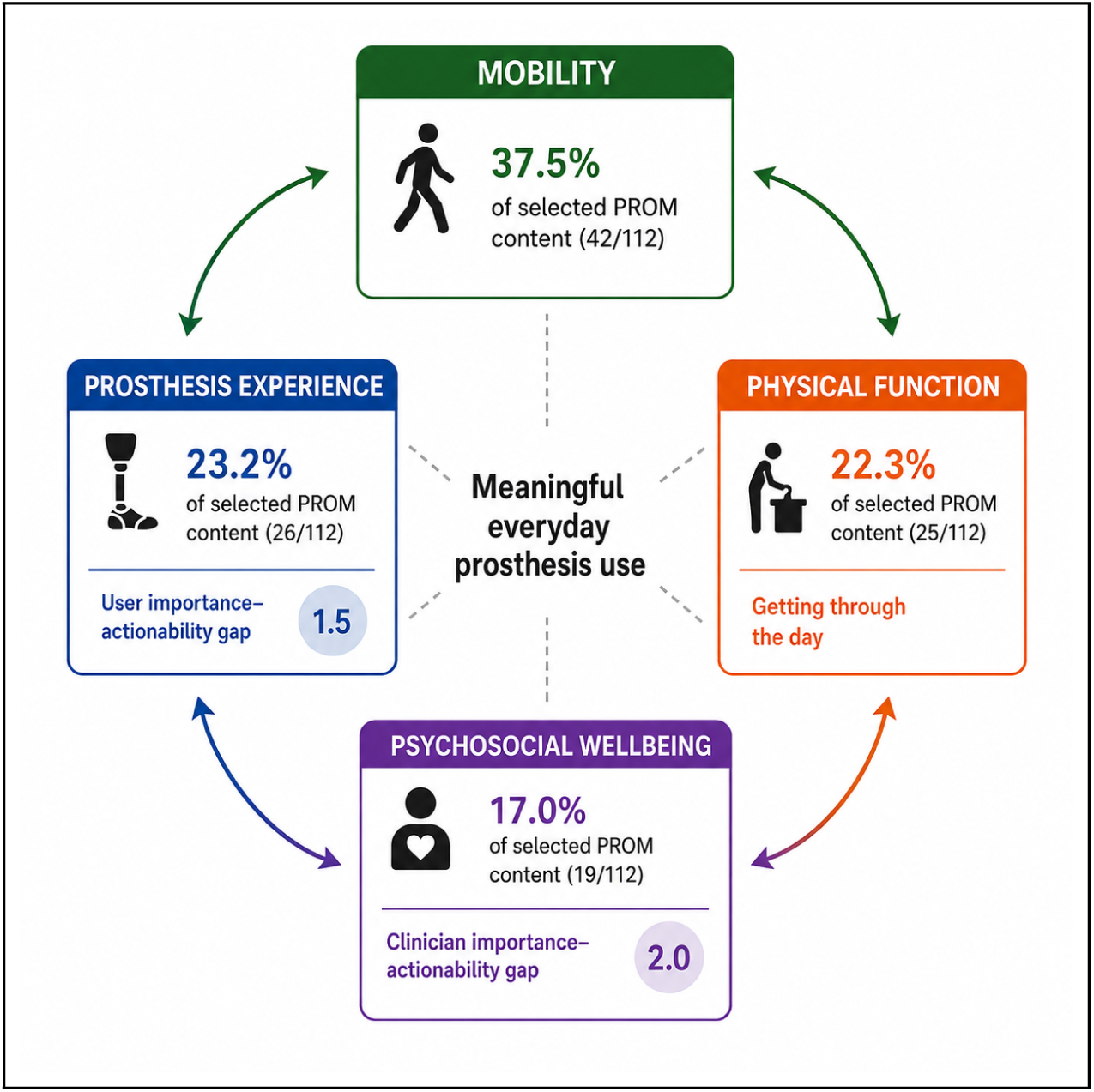
Integrated summary of the four outcome domains in meaningful everyday prosthesis use. Bidirectional arrows indicate cross-domain interdependence, while annotations summarise selected PROM-content representation and notable importance–actionability gaps.

## 4 Discussion

This study builds on prior patient-centred work identifying meaningful outcome domains following lower-limb amputation and prosthetic rehabilitation and examining patients’ experiences of outcome measurement (Ostler et al., 2023, 2024b,a). The present study extends this work through a concept-focused synthesis of purposively selected content from established PROMs and evaluation of the resulting framework with prosthesis users, clinicians, and industrial partners, enabling comparison of domain importance and actionability across stakeholder groups. The inclusion of industrial representatives provided a complementary design and commercial perspective alongside clinical and lived-experience perspectives, allowing device-related priorities to be considered within the broader interpretation of rehabilitation outcomes (Jones et al., 2023). The diversity of our sample, including participants from refugee and asylum-seeking backgrounds, varied socioeconomic circumstances, and multiple ethnic communities, also responds to longstanding concerns that rehabilitation research may underrepresent groups at greater risk of exclusion (Quigley et al., 2023).

### 4.1 Mobility

Mobility was consistently identified by contributors as a highly valued and comparatively actionable domain, representing independence, freedom of movement, and the ability to move without reliance on other people. Contributors emphasised that meaningful mobility extended beyond the capacity to walk, encompassing the confidence to move spontaneously and safely across varied and unpredictable environments. This broader interpretation is consistent with biomechanical dimensions of real-world mobility, including gait stability, dynamic balance, and movement across changing surfaces (Cutti et al., 2018; Olaya-Mira et al., 2025). Gait asymmetry in particular remains common after lower-limb amputation and is associated with slower walking speeds, although its relationship with patient-reported functional outcomes remains complex (Wong et al., 2022). Such physical capabilities also contribute to prosthetic prescription within the K-level functional classification framework, which stratifies users according to functional mobility potential and activity demands (Dillon et al., 2018). However, contributors described everyday mobility as fluctuating from day to day, influenced by socket comfort, residual-limb condition, fatigue, confidence, and environmental and social context. These lived experiences reinforce the dynamic and context-sensitive nature of mobility and suggest that performance observed under standardised conditions may not fully represent mobility as experienced in everyday life. This interpretation aligns partly with quantitative evidence. Wurdeman et al. (2018) found that self-reported mobility correlated moderately with quality of life (*r* = 0.511, *p <* 0.001) and satisfaction (*r* = 0.475, *p <* 0.001), reinforcing its broader psychosocial significance. Gailey et al. (2020) similarly identified modifiable physical factors, including hip extensor strength, hip range of motion, and single-limb balance, as significant influences on basic prosthetic mobility. Our contributors added an important contextual dimension by describing mobility and confidence as variable rather than fixed, with positive experiences supporting confidence and setbacks such as pain, discomfort, or fear of falling reducing willingness to engage in more demanding environments. Confidence may therefore be shaped by both individual factors, including pain, fatigue, and residual-limb condition, and external factors such as terrain, obstacles, social environment, and weather (Sions et al., 2020). This interpretation is consistent with evidence linking balance confidence to community participation and perceived mobility (Sions et al., 2020), as well as to the risk of subsequent injurious falls (Tobaigy et al., 2023). Earlier predictive work, including Wong et al. (2014), has demonstrated the value of balance and confidence measures in classifying prosthetic use. The present findings complement such work by suggesting that the everyday expression of confidence may vary with changing physical, environmental, and psychosocial circumstances. Physical competence and emotional security should therefore be considered as interacting influences on real-world mobility rather than as wholly separate components of rehabilitation.

### 4.2 Physical Function

Contributors described physical function as the practical capacity to sustain everyday routines rather than as a series of isolated activities. The need for, and experience of, prosthesis use varied according to the task being performed. Standing and movement-based activities more commonly required the prosthesis, whereas some seated or static activities could be completed without it. This context-dependent pattern is consistent with previous evidence that everyday prosthesis use is shaped by multiple intrinsic and extrinsic factors, including comfort and prosthetic fit (Roberts et al., 2021; Diment et al., 2022). Beyond comfort, contributors also described the practical demands associated with donning, doffing, cleaning, and adjusting the prosthesis, with burdensome preparation potentially reducing willingness to engage in everyday activities. Similar practical barriers to prosthesis use have been identified previously (Manz et al., 2022). Physical function was frequently achieved through pacing, adaptation, and energy management, with contributors placing greater value on being able to complete meaningful daily activities than on performing them quickly. This perspective suggests that successful everyday function may depend on flexible strategies that enable individuals to manage changing task demands, fatigue, and prosthesis-related constraints.

This view helps interpret evidence showing that improvements achieved during structured rehabilitation do not necessarily describe how function is subsequently experienced in everyday life. De-Rosende Celeiro et al. (2017) reported significant gains in ADL independence during structured pre-prosthetic intervention (*p <* 0.001 for most domains), while Zidarov et al. (2009) observed improvements in life-habit performance following inpatient rehabilitation alongside variation in subsequent prosthesis use. Our contributors similarly emphasised that everyday functional performance depended on continuing adaptation, comfort, pacing, and the demands of individual activities. These findings therefore highlight the importance of considering how functional capability is translated into sustained performance within everyday contexts, rather than evaluating rehabilitation success solely through performance observed during structured care.

The contributors’ emphasis on balancing effort and recovery also complements Roberts et al. (2021), who described prosthesis use as a negotiation between limitations and expectations. While Roberts focused on task-specific adjustment, our findings underline the temporal component by showing how contributors actively pace activity to preserve endurance across the day. Furthermore, the contextual variation described here echoes observations by Stuckey et al. (2020), who highlight how work and household roles interact with social expectations and fatigue. These findings suggest that the type and pattern of activities most commonly performed by a user should inform prosthesis prescription, component selection, and rehabilitation planning. In particular, ADL-specific demands such as occupational tasks, transportation use, and domestic roles should be incorporated as clinical targets alongside mobility goals (Norvell et al., 2023; Bosman et al., 2023).

### 4.3 Psychosocial Wellbeing

Psychosocial wellbeing emerged as one of the most nuanced aspects of contributors’ experiences of rehabilitation and everyday prosthesis use. Emotional adaptation, including rebuilding confidence, adjusting self-image, and learning to trust the prosthesis, was described as an ongoing process alongside physical recovery. Confidence was dynamic, strengthened through positive experiences but readily diminished by setbacks such as pain, falls, discomfort, or embarrassment in public. These accounts highlight the nonlinear nature of psychosocial adjustment and its close relationship with participation and continued prosthesis use.

Empirical studies support the relevance of these psychosocial influences. Rachmat et al. (2019) found that motivation, optimism, and social support significantly predicted subjective well-being (*p*-values between 0.02 and 0.03), while Mutar and Naji (2024) reported positive correlations between social support and self-efficacy (*r* = 0.13–0.20) in 200 users. The present findings add a temporal and contextual dimension by showing that confidence may fluctuate within everyday life in response to changing physical experiences, social situations, and prosthesis-related difficulties. This interpretation is consistent with Seti and Ned (2025), who described experiences of acceptance and withdrawal as influenced by social feedback and prosthesis comfort. Psychosocial adjustment should therefore be understood as dynamic and context-sensitive rather than represented solely by a single stable assessment.

Contributors also highlighted self-consciousness, body image, and concern about being observed or judged in public as barriers to participation. This observation is consistent with Shankar et al. (2020), whose WHOQOL-BREF findings demonstrated variation across physical, psychological, social, and environmental dimensions of quality of life among people with lower-limb amputation, with the social domain showing the lowest mean score. Importantly, the present findings showed that psychosocial difficulties could coexist with apparently adequate physical capability, reinforcing the need to consider confidence, emotional adjustment, and social participation alongside conventional measures of mobility and physical function. Rather than evolving independently of mechanical performance, these dimensions may interact with physical and prosthesis-related outcomes while also showing distinct patterns of change. Psychosocial wellbeing should therefore be considered explicitly within assessment and rehabilitation planning rather than inferred from physical performance alone.

### 4.4 Prosthesis Experience

Contributors described prosthesis experience as an evolving process shaped by trust, comfort, usability, and predictability. Comfort was consistently identified as an important influence on sustained use, affecting whether the device could be worn comfortably throughout the day or was removed when discomfort developed. When fit and function were satisfactory, contributors described greater ease and reduced conscious awareness of the device. Conversely, discomfort, heat, perspiration, skin irritation, and concerns about stability could lead to reduced activity or earlier removal of the prosthesis. These observations illustrate that prosthesis experience is not fixed, but may vary in response to changing physical, prosthesis-related, and environmental conditions (Diment et al., 2022). These lived accounts complement and deepen existing findings. Webster et al. (2012) observed that 92% of participants with dysvascular lower-limb amputation were fitted with a prosthesis by 12 months, while factors including depression, pain, age, and comorbidity were associated with aspects of prosthetic fitting, use, or functional restriction. Mohd Hawari et al. (2017) similarly reported that durability and comfort were rated as the most important socket characteristics by 83.3% of participants, while 66.7% identified socket material as the most important feature of aesthetic appearance. While our contributors echoed these priorities, they also provided an important interpretive layer by describing comfort as closely connected to trust, confidence, and willingness to engage in everyday activity. Industrial partners complemented these perspectives by emphasising usability, reliability, durability, aesthetics, and the extent to which a prosthesis could be integrated into the user’s everyday routines.

Quantitative relationships between fit and activity further illustrate this connection. Diment et al. (2022) found that more active prosthesis users wore their prosthesis for longer (*r* = 0.73) and reported greater satisfaction with socket fit (*r_s_* = 0.49), whereas community participation showed only weak associations with activity (*r_s_* = 0.13) and prosthesis comfort (*r_s_* = 0.19). This partial dissociation is consistent with the present findings, in which comfort and fit were important but interacted with confidence, psychosocial experience, and the demands of everyday participation. Although Silva et al. (2025) demonstrated that composite carbon-fibre designs can exceed mechanical standards (heel-region load 358% above ISO 10328), our contributors emphasised that technical performance alone may not fully determine how a prosthesis is experienced or used in everyday life. Trust, usability, comfort, and other experiential factors should therefore be considered alongside mechanical performance when evaluating sustained prosthesis use.

### 4.5 Environmental and Social Context

Contributors consistently described environmental and social context as important influences on when, where, and how they used their prosthesis. Confidence and mobility were often greater in familiar or predictable environments and reduced in situations involving slopes, uneven ground, poor weather, crowded public spaces, or heightened social visibility. These accounts suggest that prosthesis use and perceived independence are context-sensitive rather than determined by physical capability alone.

These experiences reinforce existing literature. Batten et al. (2020) identified terrain and public scrutiny as important barriers to community walking, while Gallagher et al. (2011) linked participation restrictions to broader contextual factors including climate, income, and infrastructure. Kam et al. (2015) similarly highlighted environmental and service-related barriers to participation in low-resource settings. The present findings extend this contextual perspective by showing that environmental and social influences remained relevant even where clinical mobility appeared adequate, particularly through their effects on confidence, safety, and willingness to participate. Environmental and social context should therefore be considered as a cross-cutting influence on mobility, physical function, psychosocial wellbeing, and prosthesis experience rather than as a separate outcome domain.

### 4.6 Integration and Broader Implications

Integration of the selected PROM-content synthesis, qualitative findings, and structured ratings supports a multidimensional interpretation of meaningful everyday prosthesis use. Mobility, Physical Function, Psychosocial Wellbeing, and Prosthesis Experience were conceptually distinct but closely interrelated, with contributors describing how changes in comfort, confidence, fatigue, environmental demands, and device performance could influence experience across more than one domain. Importantly, favourable performance within one domain did not necessarily indicate favourable outcomes in others. This was particularly evident where adequate mobility coexisted with psychosocial concerns, prosthesis-management difficulties, or restrictions in everyday participation.

The quantitative findings added an important distinction between perceived importance and perceived actionability. Mobility was consistently rated as highly important and comparatively actionable across stakeholder groups, whereas the largest importance– actionability difference among prosthesis users occurred for Prosthesis Experience (4.5 versus 3.0). Among clinicians, the largest difference occurred for Psychosocial Wellbeing (5.0 versus 3.0), indicating that clinicians considered psychosocial outcomes highly important while perceiving greater difficulty in assessing or monitoring them within regular care. The two industrial partners showed a different pattern, rating Prosthesis Experience highly for both importance and actionability. These stakeholder-specific differences suggest that perceived relevance and practical assessability are not interchangeable and may vary according to lived, clinical, and design perspectives.

This divergence between importance and actionability is consistent with broader concerns that clinical assessment may favour outcomes that are comparatively straightforward to quantify while experiential and context-sensitive aspects of rehabilitation can be more difficult to represent within routine monitoring (De-Rosende Celeiro et al., 2017; Gailey et al., 2020). Risk-prediction tools have been developed to estimate outcomes including mortality, postoperative morbidity, re-amputation, and ambulation following major lower-limb amputation; however, many lack adequate external validation, limiting their generalisability and routine clinical implementation (Preece et al., 2021). The present findings suggest that assessment of rehabilitation outcomes should therefore consider measurable physical capability alongside confidence, comfort, prosthesis experience, and the contexts in which everyday activity occurs. Rather than replacing established clinical measures, a multidimensional approach may help explain why similar levels of observed physical performance can correspond to substantially different experiences of independence and participation.

### Limitations

This study has several limitations. The stakeholder sample was relatively small and purposively recruited, which may limit transferability to wider prosthetic rehabilitation populations and may underrepresent individuals who face greater barriers to research participation (Ostler et al., 2022; Sanders et al., 2020). The PROM-content synthesis was purposive and concept-focused rather than systematic or exhaustive; therefore, the mapped content should not be interpreted as representing complete coverage of the five source instruments. The four-domain preliminary framework was developed before the main stakeholder interviews and may therefore have shaped subsequent discussion, while the importance and actionability ratings were study-specific exploratory measures rather than validated scales. Framework-evaluation interviews were audio-recorded to support accurate documentation, but full verbatim transcripts were not produced, which may have limited the depth of subsequent re-analysis. The cross-sectional design also prevented examination of how domain priorities and perceived actionability may change across the rehabilitation trajectory. Finally, clinical and industrial perspectives were drawn from relatively small groups, particularly the two industrial partners, and these findings should therefore be interpreted descriptively rather than as representative of the wider stakeholder communities.

## Conclusions

This study developed and evaluated a stakeholder-informed framework comprising four interdependent outcome domains, namely Mobility, Physical Function, Psychosocial Wellbeing, and Prosthesis Experience, that collectively reflect the complexity of everyday prosthesis use following lower-limb amputation or limb absence. The findings indicate that rehabilitation outcomes may not be fully represented by mechanical performance or isolated clinical measures alone, and that assessment should also consider emotional adaptation, prosthesis experience, and the contexts in which everyday activity occurs. Differences between observed physical performance and contributors’ lived experiences further support the value of assessment approaches that incorporate confidence, comfort, functional engagement, and real-world context alongside conventional measures of mobility. These findings provide a stakeholder-informed foundation for the further development and evaluation of outcome frameworks that are meaningful, feasible, and responsive to everyday prosthetic rehabilitation.

Future research should examine how these domains can be operationalised within routine clinical practice and evaluated longitudinally, with particular attention to developing or adapting measures that capture the dynamic and context-sensitive nature of prosthesis use over time. The participatory involvement of prosthesis users, clinicians, and industrial partners strengthened the relevance of the framework by incorporating lived, clinical, and device-development perspectives. Industrial partners contributed a complementary perspective on usability, reliability, aesthetics, and integration of the prosthesis into everyday routines, although the small industrial sample means these findings should be interpreted cautiously. Future work should therefore involve broader representation across clinical, industrial, regulatory, and policy contexts and should evaluate the framework in larger and more diverse populations.

## Data Availability

The data that support the findings of this study are not publicly available due to the sensitive and personal nature of the qualitative data collected from participants, and in accordance with the ethical approval conditions and UK GDPR obligations under which this study was conducted. Anonymised summary data may be made available upon reasonable request to the corresponding author, subject to institutional review and participant confidentiality constraints.

## Acknowledgements

The authors would like to thank all contributors who generously gave their time and shared their experiences as part of this study, including prosthesis users, clinicians, and industrial partners. Recruitment was supported by the British Association of Prosthetists and Orthotists (BAPO), whose assistance in reaching underrepresented communities was invaluable. The authors also wish to acknowledge the Amputation Rehabilitation Research Network for their support and engagement with this work. This study was conducted in accordance with the GRIPP2 reporting guidelines for PPI research (Staniszewska et al., 2017).

## Author Contributions

Mustafa Ahmed co-designed the study, developed the data collection instruments, conducted the contributor interviews, led the selected PROM-content synthesis and primary qualitative and quantitative analyses, and wrote the original draft of the manuscript. Steven Karlsson-Brown provided expertise in Patient and Public Involvement and Engagement methodology and contributed to reviewing and editing the manuscript. Pelagia Koufaki contributed to the clinical interpretation of findings and reviewed and edited the manuscript. Moghadaseh Ahmadi supported participant recruitment and data collection, contributed to the collaborative coding and mapping of selected PROM content, and reviewed and edited the manuscript. Encarna Micó-Amigo designed the study, contributed to reflexive review and interpretation of the qualitative analysis, co-wrote and reviewed the manuscript, provided supervision and project administration throughout, and acquired funding supporting this work. All authors have read and approved the final manuscript.

## Declaration of Generative AI Use

Microsoft Co-Pilot was used during manuscript preparation to assist with language refinement and structural editing. All AI-assisted content was critically reviewed, revised, and verified by the authors, who take full responsibility for the accuracy, originality, and integrity of the final manuscript. Generative AI was not used to generate, modify, or analyse the study data.

## Biographical Notes

**Mustafa Ahmed** is a Research Associate at the Institute of Mechanical, Process and Energy Engineering, Heriot-Watt University, Edinburgh. He completed his PhD in biomedical engineering at Queen Margaret University in 2026. His research focuses on wearable sensing, deep learning for activity recognition, and real-world monitoring of prosthesis use.

**Steven Karlsson-Brown** is a specialist in Patient and Public Involvement and Engagement (PPIE) at Heriot-Watt University, Edinburgh. He supports researchers in embedding meaningful public involvement across all stages of the research process and has extensive experience in co-design methodologies within health and social care research.

**Pelagia Koufaki** is a Professor of Physiotherapy at the School of Health Sciences, Queen Margaret University, Edinburgh. Her research focuses on exercise physiology, rehabilitation, and clinical assessment in chronic disease and disability populations, including individuals with limb loss.

**Moghadaseh Ahmadi** is a researcher at the School of Health Sciences, Queen Margaret University, Edinburgh. Her work focuses on prosthetic rehabilitation, outcome measurement, and the experiences of individuals with lower-limb amputation.

**Encarna Micó-Amigo** is an Assistant Professor at the Institute of Mechanical, Process and Energy Engineering, Heriot-Watt University, Edinburgh. Her research focuses on wearable sensing, real-world mobility assessment, and the development of validated digital health frameworks for rehabilitation populations.

